# Multi-population genome-wide association study implicates both immune and non-immune factors in the etiology of pediatric steroid sensitive nephrotic syndrome

**DOI:** 10.1101/2022.09.13.22279644

**Authors:** Alexandra Barry, Michelle T. McNulty, Xiaoyuan Jia, Yask Gupta, Hanna Debiec, Yang Luo, China Nagano, Tomoko Horinouchi, Seulgi Jung, Manuela Colucci, Dina F. Ahram, Adele Mitrotti, Aditi Sinha, Nynke Teeninga, Gina Jin, Shirlee Shril, Gianluca Caridi, Monica Bodria, Tze Y Lim, Rik Westland, Francesca Zanoni, Maddalena Marasa, Daniel Turudic, Mario Giordano, Loreto Gesualdo, Riccardo Magistroni, Isabella Pisani, Enrico Fiaccadori, Jana Reiterova, Silvio Maringhini, William Morello, Giovanni Montini, Patricia L. Weng, Francesco Scolari, Marijan Saraga, Velibor Tasic, Domenica Santoro, Joanna A.E. van Wijk, Danko Milošević, Yosuke Kawai, Krzysztof Kiryluk, Martin R. Pollak, Ali Gharavi, Fangmin Lin, Ana Cristina Simœs e Silva, Ruth J.F. Loos, Eimear E. Kenny, Michiel F. Schreuder, Aleksandra Zurowska, Claire Dossier, Gema Ariceta, Magdalena Drozynska-Duklas, Julien Hogan, Augustina Jankauskiene, Friedhelm Hildebrandt, Larisa Prikhodina, Kyuyoung Song, Arvind Bagga, Hae Il Cheong, Gian Marco Ghiggeri, Prayong Vachvanichsanong, Kandai Nozu, Marina Vivarelli, Soumya Raychaudhuri, Katsushi Tokunaga, Simone Sanna-Cherchi, Pierre Ronco, Kazumoto Iijima, Matthew G. Sampson

**Affiliations:** Division of Nephrology, Boston Children’s Hospital, Boston, MA, USA; Kidney Disease Initiative, Broad Institute of MIT and Harvard, Cambridge, MA, USA; Genome Medical Science Project (Toyama), National Center for Global Health and Medicine (NCGM), Tokyo, Japan; Department of Human Genetics, Graduate School of Medicine, The University of Tokyo, Tokyo, Japan; Division of Nephrology, Department of Medicine, Columbia University College of Physicians and Surgeons, New York, NY, USA; Sorbonne Université, UPMC Paris 06, Institut National de la Santé et de la Recherde Médicale, Unité Mixte de Rechereche S 1155, Paris, France; Kennedy Institute of Rheumatology, University of Oxford, Roosevelt Drive, Headington, Oxford, OX3 7FY, United Kingdom; Center for Data Sciences, Brigham and Women’s Hospital, Harvard Medical School, Boston, MA, USA; Divisions of Genetics and Rheumatology, Department of Medicine, Brigham and Women’s Hospital, Harvard Medical School, Boston, MA, USA; Program in Medical and Population Genetics, Broad Institute of MIT and Harvard, Cambridge, MA, USA; Department of Pediatrics, Kobe University Graduate School of Medicine, Kobe, Japan; Department of Biochemistry and Molecular Biology, University of Ulsan College of Medicine, Songpa-gu, Seoul, Korea; Renal Diseases Research Unit, Genetics and Rare Diseases Research Division, Istituto di Ricovero e Cura a Carattere Scientifico Ospedale Pediatrico Bambino Gesù, Rome, Italy; Nephrology, Dialysis and Transplantation Unit, Department of Emergency and Organ Transplantation, University of Bari Aldo Moro, Bari, Italy; Department of Pediatrics, AIIMS, New Delhi, India; Department of Pediatric Nephrology, Amalia Children’s Hospital, Radboud University Medical Center, Nijmegen, The Netherlands; Department of Medicine, Boston Children’s Hospital, Boston, MA, USA; Department of Pediatrics, Harvard Medical School, Boston, MA, USA; Laboratory on Molecular Nephrology, IRCCS Instituto Giannina Gaslini, Genoa, Italy; Department of Nephrology and Renal Transplantation, IRCCS Instituto Giannina Gaslini, Genoa, Italy; Department of Pediatric Nephrology, VU University Medical Center, Amsterdam, The Netherlands; Division of Transplantation, Department of Surgery, University of Pennsylvania, Philadelphia, PA; Department of Pediatric Nephrology, Dialysis and Transplantation, Clinical Hospital Hospital Center Zagreb, University of Zagreb Medical School, Zagreb, Croatia; Division of Nephrology and Pediatric Dialysis, Bari Polyclinic Giovanni XXIII Children’s Hospital, Bari, Italy; Department of Nephrology, Dialysis and Transplant Unit, University Hospital of Modena, Modena, Italy; Surgical, Medical and Dental Department of Morphological Sciences, Section of Nephrology, University of Modena and Reggio Emilia, Modena, Italy; Unità Operativa Nefrologia, Azienda Ospedaliero-Universitaria di Parma, Dipartimento di Medicina e Chirurgia, Università di Parma, Parma, Italy; Department of Nephrology, Medicine and General University Hospital, Charles University, Prague, Czech Republic; Department of Pediatrics, ISMETT, Palermo, Italy; Pediatric Nephrology, Dialysis and Transplant Unit, Fondazione IRCCS Ca’ Granda-Ospedale Maggiore Policlinico, Milano, Italy; Department of Clinical Sciences and Community Health, University of Milan, Italy; Department of Pediatric Nephrology, UCLA Medical Center and UCLA Medical Center-Santa Monica, Los Angeles, CA, USA; Department of Medical and Surgical Specialties, Radiological Sciences, and Public Health, Division of Nephrology and Dialysis, University of Brescia and ASST Spedali Civili of Brescia, Brescia, Italy; Department of Pediatrics, University of Split, Split, Croatia; Department of Pediatric Nephrology, University Children’s Hospital, Skopje, Macedonia; Division of Nephrology and Dialysis Unit, University of Messina, Sicily, Italy; Division of Nephrology, Beth Israel Deaconess Medical Center, Boston, MA, USA; Department of Pediatric, Division of Pediatric Nephrology, Columbia University Irving Medical Center New York-Presbyterian Morgan Stanley Children’s Hospital in New York, NY, USA; Department of Pediatrics, Interdisciplinary Laboratory of Medical Investigation, Faculty of Medicine, Federal University of Minas Gerais, Belo Horizonte, Brazil; The Charles Bronfman Institute for Personalized Medicine, Icahn School of Medicine at Mount Sinai, New York, NY, USA; Institute for Genomic Health, Icahn School of Medicine at Mount Sinai, New York, NY, USA; Division of Genomic Medicine, Department of Medicine, Icahn School of Medicine at Mount Sinai, New York, NY, USA; Division of General Internal Medicine, Department of Medicine, Icahn School of Medicine at Mount Sinai, New York, NY, USA; Department of Pediatrics, Nephrology and Hypertension, Medical University Gdansk, Poland; AP-HP, Pediatric Nephrology Department, Hôpital Robert-Debré, Paris, France; Pediatric Nephrology, Hospital Universitari Vall d’Hebron, Universitat Autónoma de Barcelona, Barcelona, Spain; Institute of Clinical Medicine, Faculty of Medicine, Vilnius University, Vilnius, Lithuania; Research and Clinical Institute for Pediatrics, Pirogov Russian National Research Medical University, Taldomskava St., 2, Moscow, Russia; Department of Pediatrics, Hallym University Sacred Heart Hospital, 22, Gwanpyeong-ro 170 beon-gil, Dongan-gu, Anyang-si, Gyeonggi-do, 14068 Korea; Department of Pediatrics, Faculty of Medicine, Prince of Songkla University, Hat-Yai, Songkhla 90110, Thailand; Division of Nephrology, and Dialysis, Department of Pediatric Subspecialities, Istituto di Ricovero e Cura a Carattere Scientifico Ospedale Pediatrico Bambino Gesù, Rome, Italy; Department of Biomedical Informatics, Harvard Medical School, Boston, MA, USA; Centre for Genetics and Genomics Versus Arthritis, University of Manchester, Manchester, UK; Department of Nephrology, Centre Hospitalier du Mans, Le Mans, France; Hyogo Prefectural Kobe Children’s Hospital, Kobe, Japan; Department of Advanced Pediatric Medicine, Kobe University Graduate School of Medicine, Kobe, Japan

## Abstract

Pediatric steroid-sensitive nephrotic syndrome (pSSNS) is the most common childhood glomerular disease. Previous genome-wide association studies (GWAS) identified a risk locus in the HLA Class II region and three additional signals. But the genetic architecture of pSSNS, and its genetically driven pathobiology, is largely unknown. We conducted a multi-population GWAS meta-analysis in 38,463 participants (2,440 cases) and population specific GWAS, discovering twelve significant associations (eight novel). Fine-mapping implicated specific amino acid haplotypes in HLA-DQA1 and HLA-DQB1 driving the HLA Class II risk signal. Non-HLA loci colocalized with eQTLs of monocytes and numerous T-cell subsets in independent datasets. Colocalization with kidney eQTLs was lacking, but overlap with kidney cell open chromatin suggests an uncharacterized disease mechanism in kidney cells. A polygenic risk score (PRS) associated with earlier disease onset in two independent cohorts. Altogether, these discoveries expand our knowledge of pSSNS genetic architecture across populations and provide cellspecific insights into its molecular drivers.

---

pSSNS is a rare disease of glomerular filtration barrier failure. Its incidence ranges from 0.96-13.5/100,000, being most frequent in South Asian and East Asian populations^1^. pSSNS causes massive proteinuria and increased risk of thromboembolism, sepsis, and progression to chronic kidney disease (CKD)/end-stage kidney disease (ESKD)^2–7^. And those progressing to ESKD have increased odds of recurrent NS in their allograft^8^. pSSNS is impactful across the lifespan - 31-50% of those affected have relapses in adulthood^9^. Much of pSSNS’s morbidity is related to side effects of the non-specific immunosuppressants that allow some to achieve remission of their proteinuria^7,10–17^.

There are no monogenic forms of pSSNS to illuminate its pathobiology. However, we know that immune dysregulation is a major contributor^18,19^. But determining causal immune factors via case-control studies of cytokines profiles, cell types, and transcriptomic signatures is challenging. The dynamic responses of the immune system at different disease stages and to various stimuli make it difficult to determine whether observed differences are causal, correlated, or due to independent biological/environmental factors. And kidney tissue in children is rarely available to determine intrarenal, molecular drivers of pSSNS.

GWAS have discovered four pSSNS risk loci^20–24^. The top signal in each was in the HLA Class II region. Two other loci are plausibly immune-related, with the closest genes being Calcium Homeostasis Modulator Family Member 6 (*CALHM6*)^25^ and TNF Superfamily Member 15 (*TNFSF15)*. The lead SNP of the fourth locus was within nephrin (*NPHS1)*, a fundamental glomerular gene implicated in Mendelian NS^26^. These studies are illuminating but naturally by smaller sample sizes, primarily population-specific analyses, and limited post-GWAS analysis. Here, towards discovering a fuller spectrum of disease-associated genetic variation and unraveling its pathogenesis at the interface of the immune system and kidney, we conducted a large and diverse GWAS of pSSNS.

We conducted a multi-population, fixed-effect, inverse-variance, meta-analysis across twelve GWAS datasets comprised of 2,440 cases and 36,023 controls of Admixed American, African, East Asian, European, Maghrebian and South Asian populations (**Figure 1, Figure S1, Table S1**).To account for population-driven effect heterogeneity, we also performed a meta-regression with MR-MEGA^27^. Eight loci (four new, and all outside HLA) were significant (MR-MEGA p < 5 × 10^−8^) (**Table 1, Figure 2A, Figure S2**). The lead SNPs of the novel loci were all intronic: **(1)** rs7759971 in Abelson Helper Integration Site 1*(AHI1*; p = 4.90 × 10^−12^); **(2)** rs55730955 in CD28 molecule (*CD28*; p = 4.27 × 10^−10^); **(3)** rs8062322 in C-type Lectin Domain Containing 16A (*CLEC16A*; p = 1.61 × 10^−10^); **(4)** rs28862935 in betacellulin (*BTC*; p = 1.08 × 10^−9^). Four other significant loci were previously reported^24,23^. Two more significant loci emerged after conditioning: **(5)** rs1794497 upstream of *HLA-DQB1*, (p = 6.79 × 10^−52^); **(6)** rs2256318 in an intron of MHC Class I Chain-related Gene A (*MICA;* p = 9.70 × 10^−18^) (**Figure 2B, Figure S3**). Population specific GWAS meta-analysis discovered two additional significant loci in Europeans (**Figure 2C, Table S2, Table S3, Figure S4**): The lead SNPs were in introns of **(7)** rs111796602 in an intron of Engulfment and Cell Motility 1 (*ELMO1;* p = 1.72 × 10^−8^) and **(8)** rs12911841 in an intron of Mortality Factor 4 Like 1 (*MORF4L1*; p = 3.88 × 10^−8^). The *CALHM6* association appeared specific to Europeans *(P*_anc_het_ *=* 4.99 × 10^−4^) and the *TNFSF15* and *NPHS1* associations to East Asians *(P*_anc_het_ *=* 7.76 × 10^−4^ and *P*_anc_het_ *=* 2.43 10^−4^, respectively) (**Figure S5, Figure S6**). All other loci had similar effects across populations (**Figure 2D**). Finally, there were 13 novel suggestive loci (MR-MEGA p < 5 × 10^−6^) in the multi-population GWAS (**Table S4, Table S5**). On a liability scale and excluding HLA, European heritability was 0.04 [CI: -0.08,0.16] and East Asian heritability was 0.12 [CI: 0.04,0.21], with large confidence intervals likely due to small effective sample sizes.

**Table 1.**
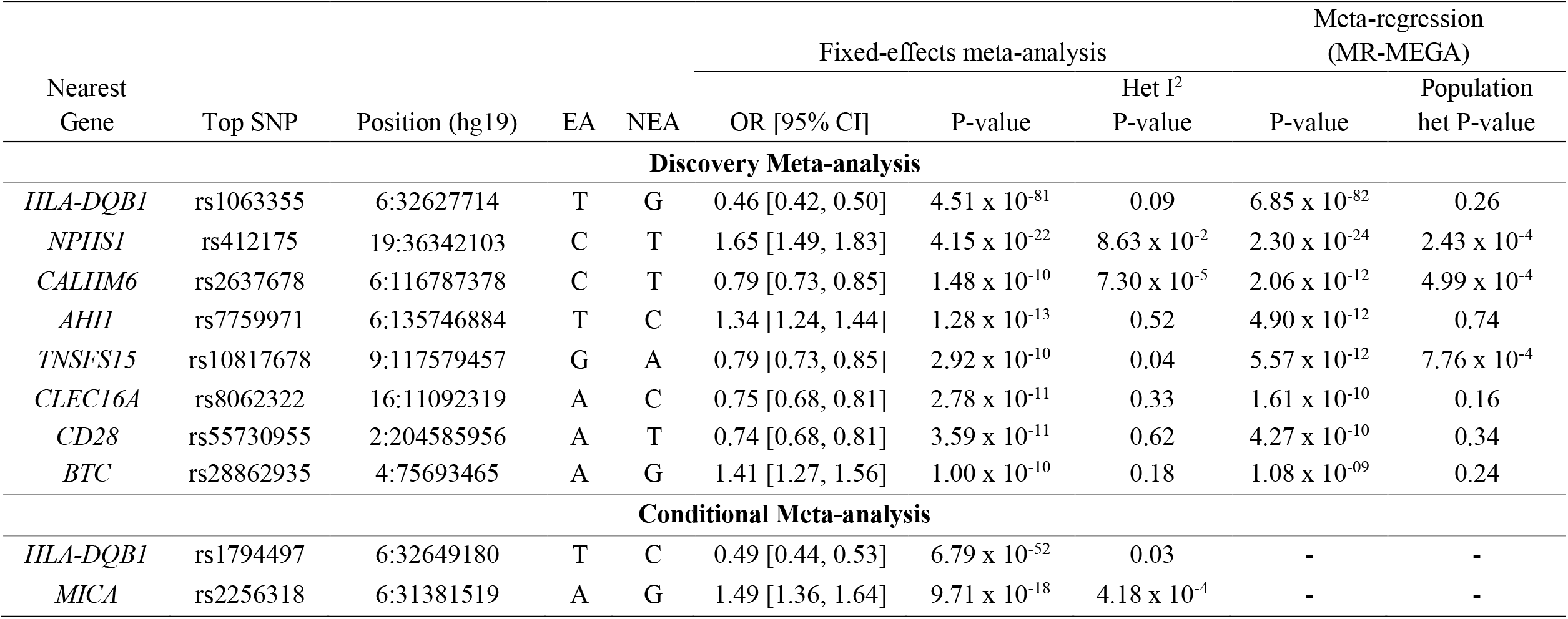
Genome-wide significant SNPs from multi-population meta-analysis. EA=effect allele, NEA=non-effect allele, OR [95% CI]= Odds ratio with 95% confidence interval. MR-MEGA results are not available for the conditional analysis.

**Figure 1:**
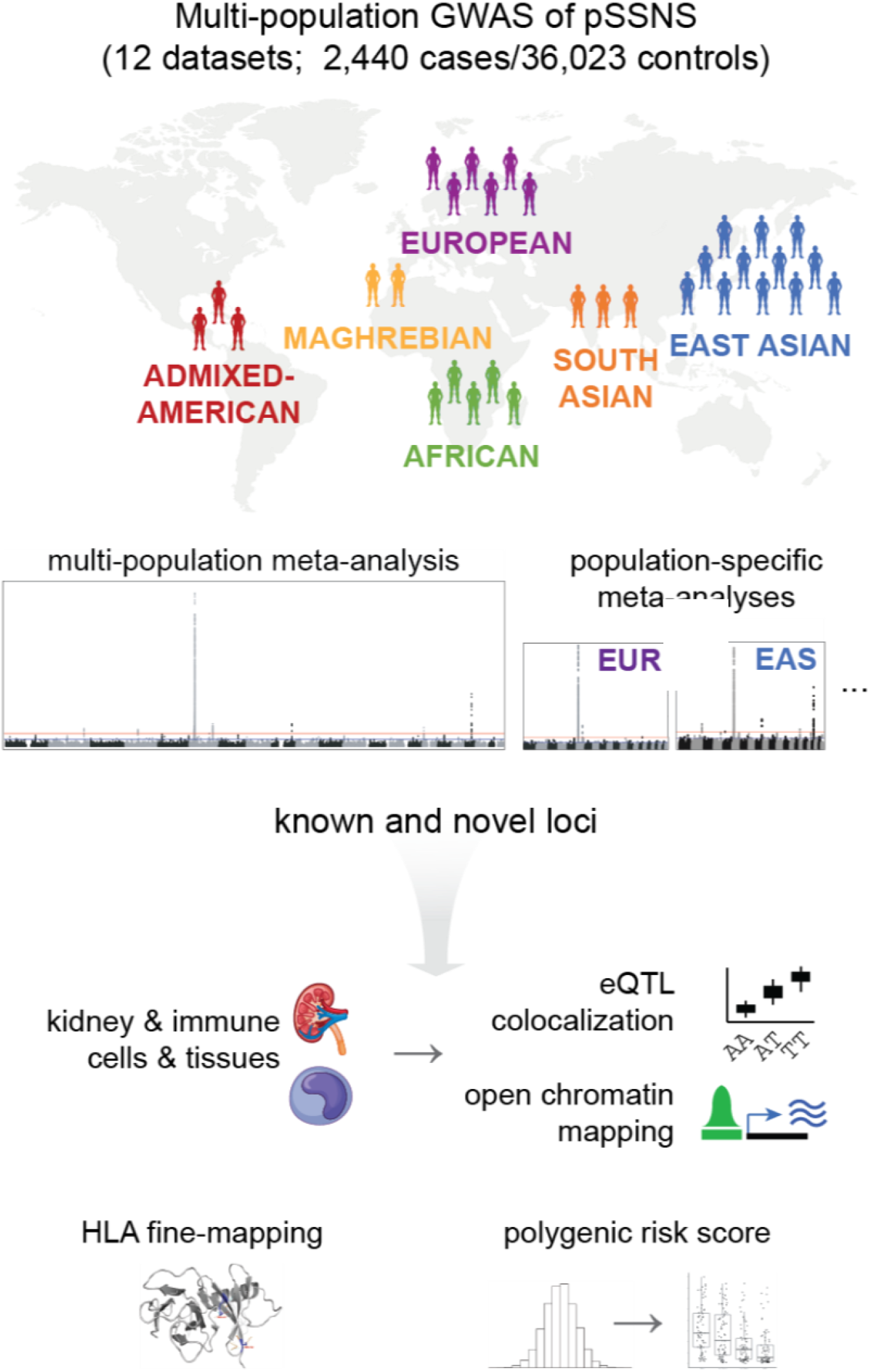
Flowchart of study design. pSSNS= pediatric steroid sensitive nephrotic syndrome, EUR = European, EAS = East Asian, eQTL = expression quantitative trait loci.

**Figure 2.**
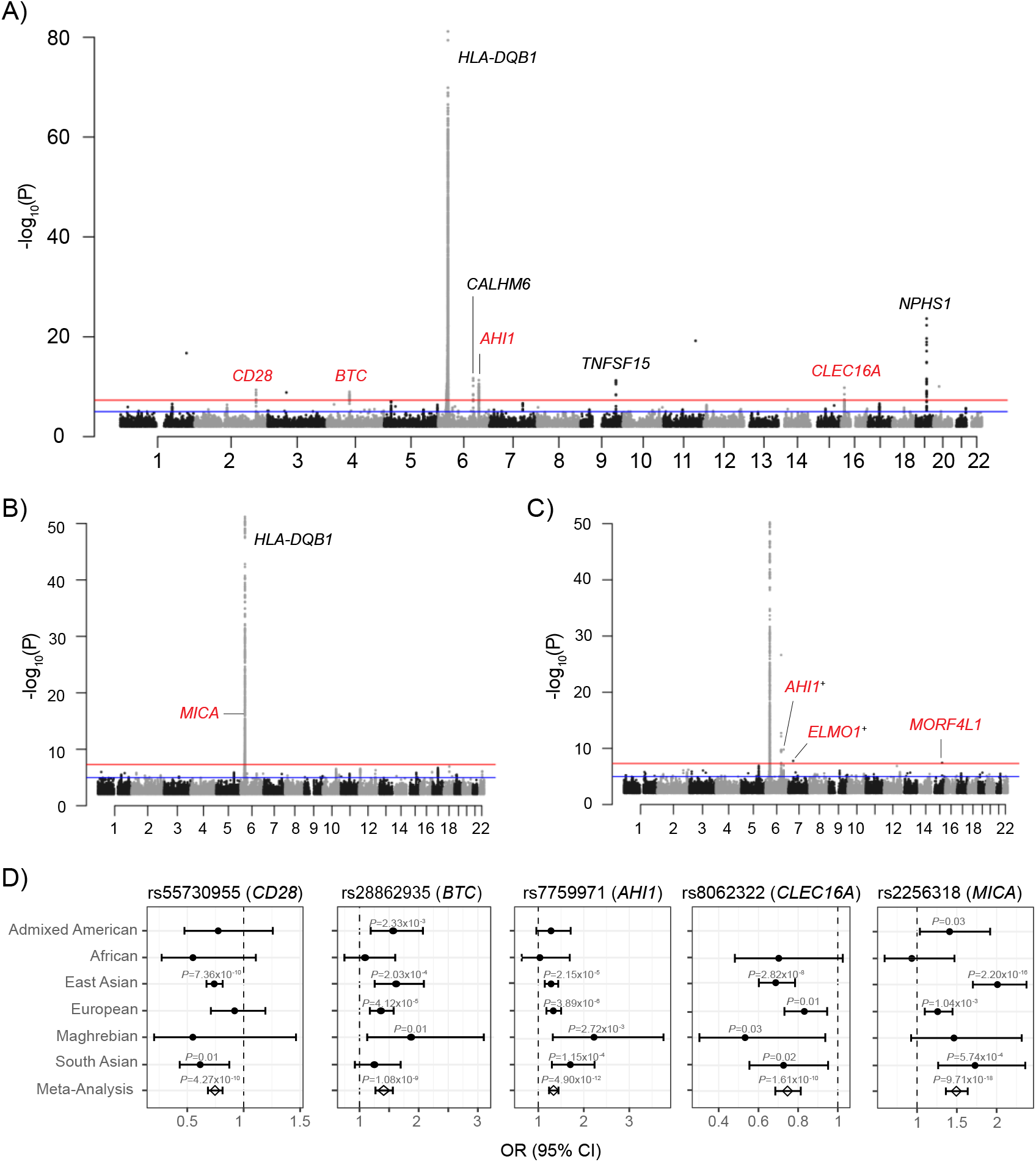
Manhattan plots of **A)** multi-population meta-analysis of 2,440 cases vs. 36,023 controls, **B)** multi-population conditional meta-analysis, **C)** European meta-analysis of 674 cases vs. 6,817 controls. Novel genome-wide association are indicated with in red and only novel associations are labeled in B and C. Discoveries that included the summary statistics from suggestive SNPs available from Dufek et al indicated with ‘+’, **D)** Multi-population & single population odds ratios for novel multi-population significant SNPs discovered

A number of insights emerged from evaluating disease associations, functions, and expression patterns of the lead SNPs and/or the closest genes at the novel non-HLA loci. First, PheWAS using Open Target Genetics (http://genetics.opentargets.org)^28^ found that SNPs at most loci associated with GWAS of diverse white blood cell traits, atopic disorders, and autoimmune conditions. For example, among the strongest associations with the lead SNPs at the following loci include: *CLEC16A, CD28, MICA*, and *ELMO1* with eosinophil counts; *AHI* with monocyte and neutrophil counts, asthma, and hay fever (also shared by *CD28*); and *MICA* with Type 1 Diabetes (T1DM).

Second, most of these genes, while primarily known for their role in immunity, also have known roles in kidney diseases and cells. Common *AHI1* variants are associated with atopy, lupus, and diverse immune cell traits^28^. Rare *AHI1* coding variants cause the monogenic ciliopathy Joubert Syndrome, which includes cystic kidney disease^29^. *ELMO1* participates in Rac1 pathway activation and actin cytoskeletal rearrangement^30^, is expressed in podocytes^31^, and is associated with diabetic nephropathy^32^. *CD28*, a T-cell glycoprotein, binds a co-stimulatory molecule B7-1 (CD80) on antigen-presenting cells. B7-1 is expressed on human podocytes in some forms of nephrotic syndrome, and blocking the B7-1/CD28 interaction with a CTLA-4 immunoglobulin can ameliorate proteinuria^33^. *MICA* is expressed in kidney endothelium, binds and activates cytotoxic CD8+ T cells and NK cells, and has increased glomerular expression in lupus^34^. *BTC* contributes to inflammation by binding to epidermal growth factor receptor^35^, a gene whose renal expression is upregulated after kidney injury^36^. *CLEC16A* takes part in the B cell receptor-dependent HLA-II pathway in human B cells^37^, but is also significantly expressed in the human podocytes (https://atlas.kpmp.org). It is involved in autophagy, mitophagy, and endolysosomal trafficking in multiple cell types^38,39^. It is also in close proximity to *CIITA*, a master transcription factor of HLA class II genes^40^ and Dexamethasone Inducible Transcript (*DEXI)*, a glucocorticoid-induced gene^41^.

We next turned to discovering specific variants and genes driving these GWAS signals and discerning whether they are acting in immune cells, kidney cells, or both.

First, we conducted colocalization with eQTL data from two functionally distinct kidney compartments (glomerulus and tubulointerstitium)^42^, multiple tissues from GTEx^43^, and immune cells from DICE^44^ and Blueprint^45^. Overall, pSSNS GWAS SNPs demonstrated significant enrichment in multiple immune cell eQTLs, led by a 69x and 62x increased odds of being monocyte and CD4+ memory Treg eQTLs, respectively (**Figure 3**). On an individual gene level, considering eQTLs with a regional colocalization probability (RCP) > 0.2, nine genes – including three which were the closest gene to a GWAS SNP (*CALHM6, AHI1, TNFSF15) –* colocalized with immune cell eQTLs (Figure 3, Table S6). All three significantly colocalized with monocyte eQTLs. *AHI* had significant eQTLs across monocytes, many T-cell subsets, and naïve B cells. Finally, in CD4+ memory Treg cells, SNPs in the “Gasdermin B (*GSDMB)”* suggestive locus colocalized with two different genes – *GSDMB* and ORMDL sphingolipid biosynthesis regulator 3 (*ORMDL3)*. The *GSDMB/ORMDL3* locus is associated with multiple autoimmune disorders and eosinophilic inflammation-driven asthma^46^. In asthma, higher *GSDMB* expression is correlated with increased interferon signaling and MHC class I antigen presentation^47^. Notably, there was no colocalization with kidney eQTLs despite sufficient sample sizes to do so (**Figure S7**).

**Figure 3.**
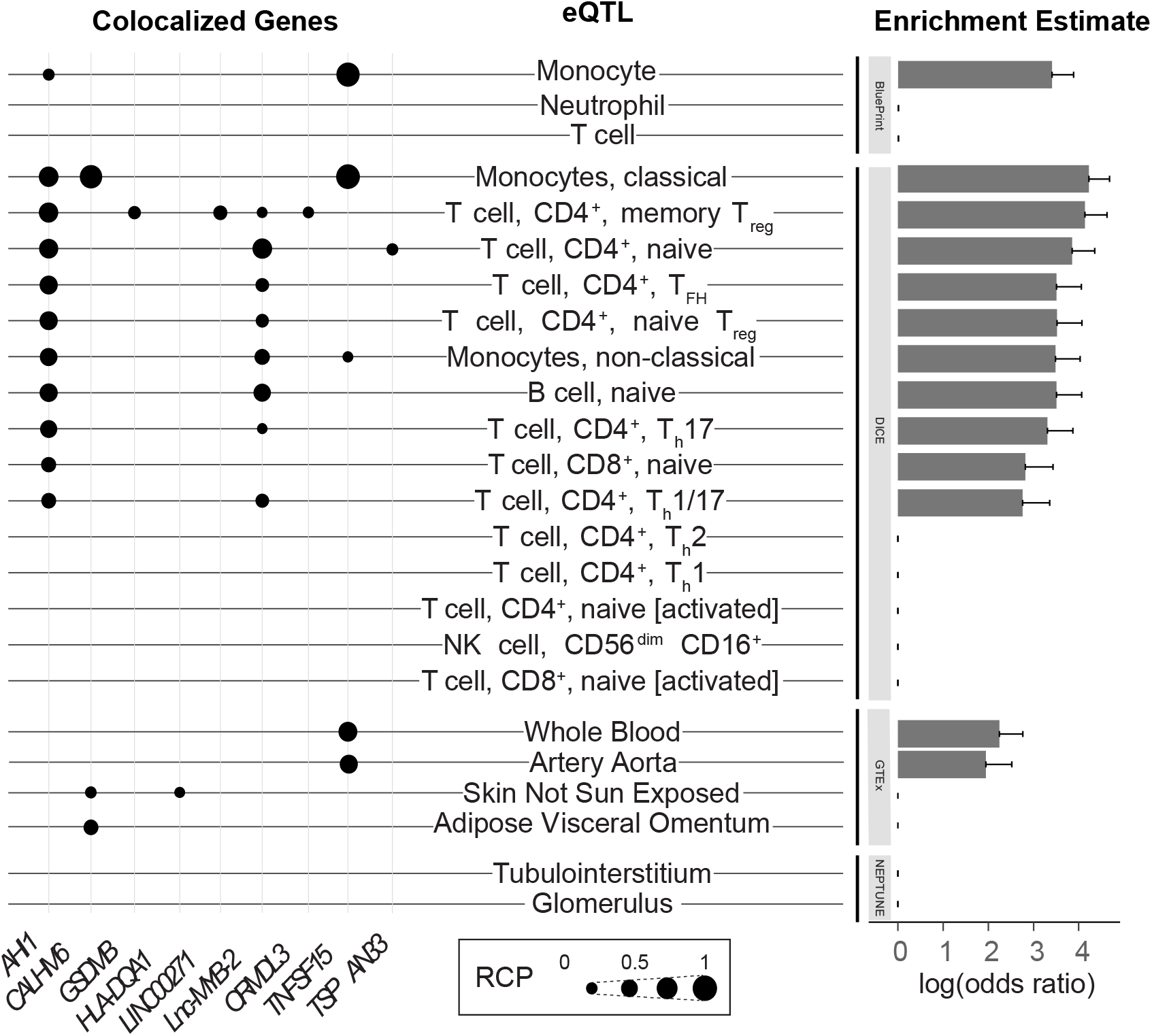
Colocalization of SSNS GWAS and eQTL datasets. Each eQTL data set is labeled with colocalized loci to the left and enrichment estimates to the right. Genes with regional colocalization probability (RCP) > 0.2 in at least one tissue/cell are included. pSSNS GWAS loci that colocalized with tissue/cell-type eQTLs are indicated by black dots, with larger dots indicating higher RCP. GTEx tissues without associations are excluded from this figure (see Figure S7). Enrichment estimates, from fastENLOC, are based on genome-wide summary statistics from GWAS and include a shrinkage parameter, resulting in 0 enrichment for multiple tissues/cell-types. **logOR=2 ∼ OR=7.5, logOR=3 ∼ OR=20.1, logOR=4 ∼ OR=50.6**.

We then created a 95% credible set for all non-HLA significant loci and assessed their overlap with ATAC-seq derived open chromatin data from immune^48^ and kidney cells^49,50^ (**Table S7**). The SNPs with the highest posterior inclusion probability (PIP) for *AHI*, rs7759971 (PIP 0.52), overlapped with open chromatin of multiple immune cell types, including CD34+ cells, common lymphoid and myeloid progenitors, hematopoietic stem cells, and multipotent progenitors. The top PIP SNP for *CLEC16A, NPHS1, CD28, CALHM6*, and *TNFSF15* had no overlap with open chromatin. However, each locus had individual SNPs with lower PIPs that overlapped with both immune and kidney cell open chromatin.

We next fine-mapped the HLA signal (**Table S8**) We first imputed across the extended MHC region using a multi-population HLA imputation panel ^51^, resulting in 640 classical HLA alleles, 4,513 amino acids in HLA proteins and 49,321 SNPs in the extended MHC region for association. We used population-specific and multi-population SNP-level logistic regression, to identify specific SNPs and classical alleles associated with pSSNS (**Table S9, Supplement Note 1, Table S10, Figure S8**).

We next turned to discovering specific HLA amino acid positions most associated with risk of pSSNS through logistic regression analysis of all residues at each position. Amino acid position 47 in *HLA-DQA1* was the most strongly associated with pSSNS (*P*_omnibus_ = 7.73 × 10^−83^) (**Table S11, Figure S9**). Arginine was the most frequent amino acid; a substitution to lysine conferred the greatest disease risk (*P* = 5.70 × 10^−80^; OR [95% CI] =3.62 [3.17 – 4.14]). A second association in near-perfect linkage disequilibrium was identified at *HLA-DQA1* position 52 (*P* = 1.14 × 10^−82^). Arginine was again the most common amino acid at this position, and a substitution to serine conferred the greatest protection from risk (*P* = 1.00 × 10^−28^; OR = 0.53 [0.47 – 0.59]). After conditioning, an independent association was discovered at *HLA-DQB1* position 26 (*P* = 3.22 × 10^−13^). A change from the most common amino acid leucine to glycine conferred the most significant protection (*P* = 4.75 × 10^−12^; OR = 0.64 [0.60 – 0.73]). A haplotype analysis identified the 47_lysine_-52_histidine_ haplotype associated with greatest increased odds of pSSNS (**Figure 4a**). *HLA-DQA1* position 47 is located on the outside of the peptide-binding groove and acts as a regulator of binding stability, which, when altered, has been suggested to mediate the development of autoimmune disorders^52^. Arginine at *HLA-DQA1* position 52 has been associated with autoimmune disorders, including T1DM^53^.

**Figure 4.**
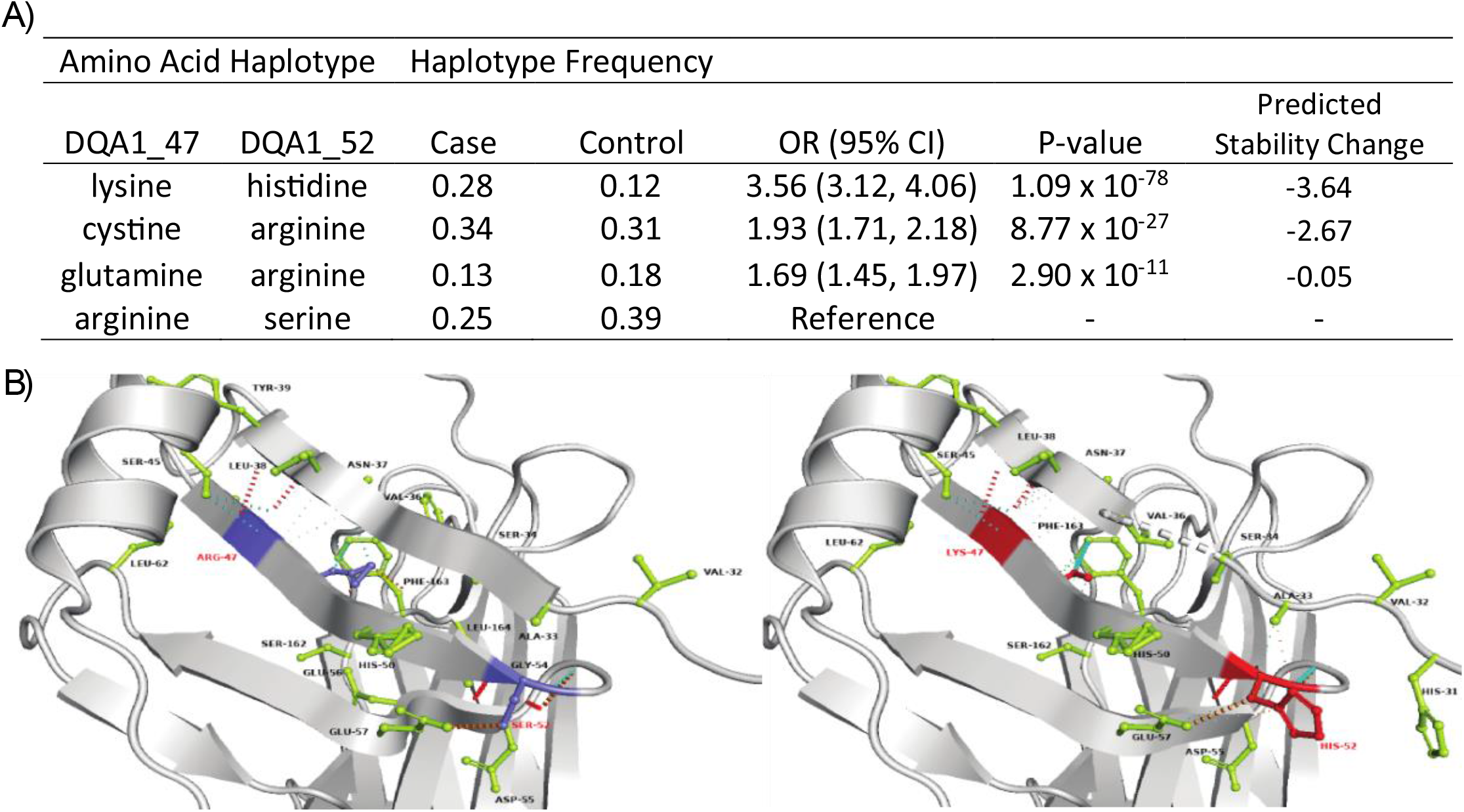
*HLA-DQA1* amino acid risk associations. **A)** Increased risk and predicted stability change of the two-amino acid residue hapltoypes at *HLA-DQA1* positions 47 and 52. Odds ratios are from a joint logistic regression with arginine_47_-serine_52_. The reference haplytype is the one conferring the strongest protection (i.e., odds ratio indicate increase in risk compared to arginine_47_-serine_52_). Increasingly negative “Predicted Stability Change” indicates increasingly decreased stability. **B)** Protein structure for the reference haplotype arginine_47_-serine_52_ (left, blue) and lysine_47_-histidine_52_ (right, red). The residues in lime color displays potential interacting amino acid with mutated amino acids. The color scheme for interaction is as follows: Cyan for Van der Waals, red for hydrogen bonds, green for hydrophobic bonds, sky blue for Carbonyl bonds and orange for polar bonds. When no bonds are displayed but the amino acids are shown they were predicted to form weak VDW bonds.

We then used DynaMut2^54^ to model the impact on protein structural stability of two amino acid haplotypes (**Figure 4b**). This is quantified by Delta Delta G (ddG), where ddG < 0 predicts unstable structure. The haplotype consisting of lysine (47) and histidine (52) predicted the most instability (ddG = -3.64). Notably, the predicted increase in protein instability and increased odds of disease for each haplotype were concordant. This suggests that pSSNS-associated haplotypes increased the odds of disease by increasing the instability of *HLA-DQA1* and altering its ability to properly form a stable HLA-II molecule.

Finally, we generated a pSSNS PRS using summary statistics of 1,607 cases and 11,995 controls from European and East Asian populations. We first tested its association with demographic and clinical phenotypes in 233 European children with sufficient clinical data from the EU-European sub-cohort, after adjusting for genetic principal components. There was a significant negative association observed between PRS and age of disease onset (*P* = 1.49 × 10^−4^, **Figure 5A, Table S12**). We then assessed the PRS in an independent cohort of 165 children with proteinuric kidney disease enrolled in the Nephrotic Syndrome Study Network (NEPTUNE)^55^. 30%, 49%, and 21% of all participants had focal segmental glomerulosclerosis, minimal change disease, or no biopsy, respectively. Adjusting for sex, histology and genetic principal components, we found a significant association between PRS and age of onset (*P* = 0.003; **Figure 5B, Table S12, Figure S10**).

**Figure 5.**
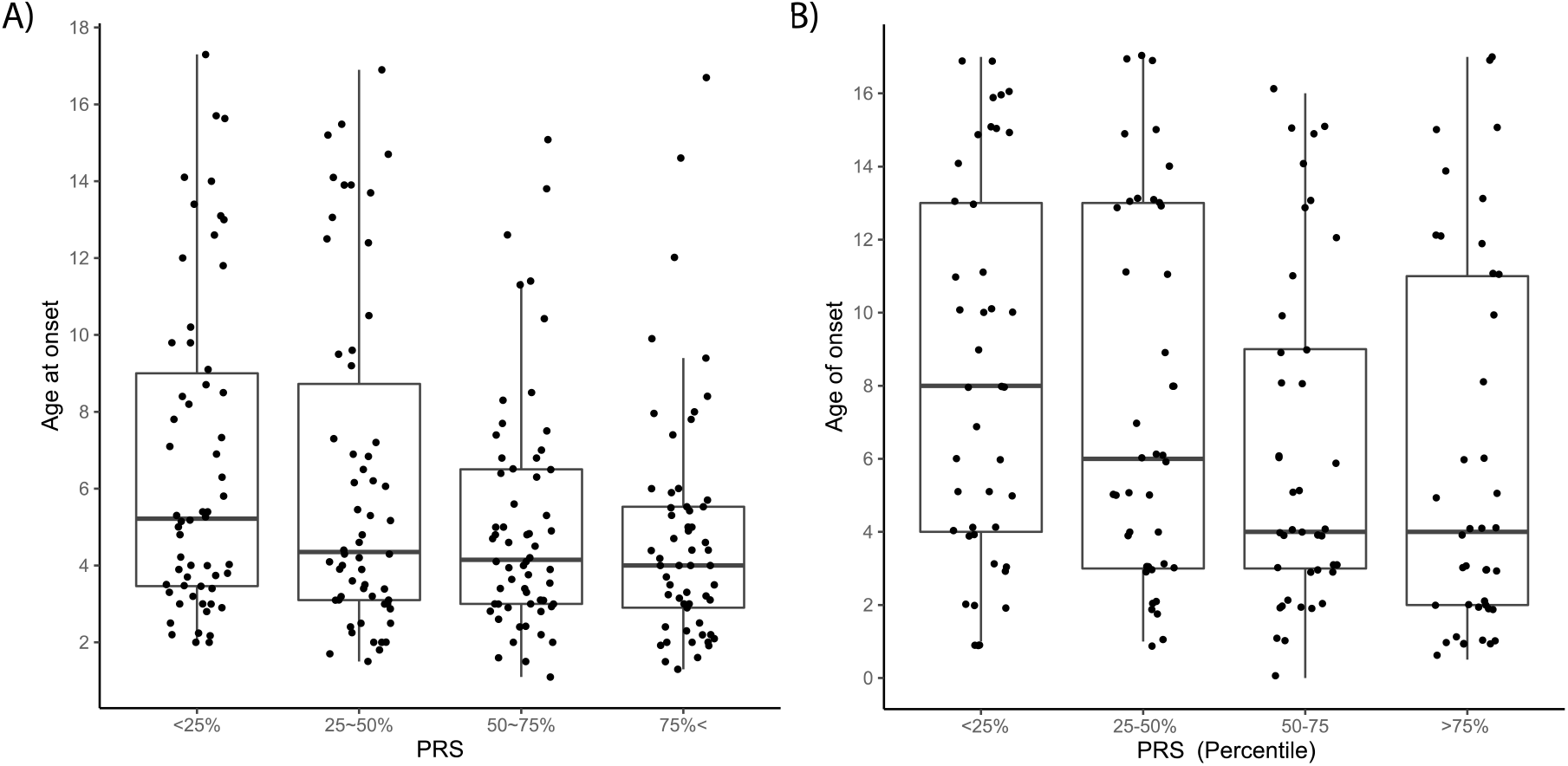
Association between pSSNS PRS quartiles and age of onset in pediatric SSNS patients from **A)** EU-European and **B)** NEPTUNE pediatric cohorts.

A number of important discoveries emerged from this study. First, we tripled the number of pSSNS from four to twelve. Second, we found that while the immunological connections with the lead SNPs and closest genes in these newly discovered loci are well-established, most of them also have a *bona fide*, but overall less understood role, in kidney cells and diseases. Discovering upon which cells and organ systems each of these risk loci act will be an important future step. The availability of omics data from pediatric kidney tissue will be critical, as we hypothesize that the paucity of kidney eQTL signals we observed may be due to mapping pSSNS GWAS data to signatures primarily from adult tissues. Third, we identified monocytes and eosinophils dysfunction as a potential contributor to the pathogenesis of pSSNS. The mechanisms by which genetically driven changes in these cell types contribute to pSSNS onset is an important area of future inquiry. Fourth, we discovered specific amino acid changes in HLA-DQA1 and -DQB1 associated with pSSNS that should help will empower subsequent studies to illuminate pathomechanisms at the most significant risk locus for this disease. Finally, the association of higher PRS with younger age of onset suggests that a stronger genetic predisposition to disease lowers the threshold of an individual to develop pSSNS in the context of environmental factors and may ultimately help share clinical screening and care. In conclusion, these findings expand our knowledge of the genetic architecture of pSSNS and accelerate our understanding of its molecular underpinnings and clinical implications.

## METHODS

### GWAS data summary

#### GWAS data from NEPHROVIR/EU

Sample collection and genotype calling were done at Sorbonne Université in Paris. Pediatric steroid sensitive nephrotic syndrome was defined as proteinuria > 0.25g/mmol, serum albumin <25 g/L (<30 in France), full response within 4 weeks of 60 mg/m^2^/day of oral prednisone or prednisolone, and age of onset < 18 years old. 244 previously reported European patients from the NEPHROVIR study^21^ were combined with 159 newly recruited European patients recruited from France, Lithuania, Poland, Russia, Italy. Healthy adult controls (n=300) were recruited from Lyon, France, and combined with population-matched controls from The 1000 Genomes Project Phase 3 release (n=493)^56^. There were also 56 sub-Saharan African cases with 454 population-matched controls from The 1000 Genomes Project and 85 Maghrebian cases with 261 Moroccan population-matched controls. Both were reanalyzed from a previous report^21^. There were 160 Indian cases with 93 population-matched controls. Samples were genotyped on the Illumina Human OmniExpress or Illumina Omni 2.5 arrays.

#### GWAS data from Columbia University (US Cohorts)

Sample collection and genotype calling was done at Columbia University in New York. Cases were defined by local recruitment centers across the US, Europe, and Brazil as either minimal change disease or non-biopsied SSNS with age of disease onset <21. Five cohorts from Columbia University consisted of patients from European (n_cases_=371, n_controls_-=4359), East Asian (n_cases_=17, n_controls_=443), sub-Saharan African (n_cases_=65, n_controls_=7344), South Asian (n_cases_ =39, n_controls_=534) and Admixed American (n_cases_ =109, n_controls_=13266) populations. The genotyping of the cases was performed using multiple versions of MEGA (Multi-Ethnic Global Array) chips that includes MEGA 1.0, MEGA 1.1 and MEGA^EX^. The controls that were genotyped on MEGA1.0 were downloaded from NCBI dbGAP (IDAT files) from the PAGE consortium^57^. The differences between the chips were corrected first by mapping all the SNPs to a common cluster file in Genome Studio for individual MEGA platforms and further using Snpflip (https://github.com/biocore-ntnu/snpflip) software.

#### GWAS data from Kobe University

Pediatric steroid sensitive nephrotic syndrome cases were defined as urine protein to creatinine ratio ≥ 2.0, serum albumin ≤ 2.5 g/dl, and complete remission with 4-6 weeks after starting 6-mg/m1^2^ oral prednisolone per day and age of onset < 18 years old. Three GWAS studies of SSNS in Japanese (n_cases_= 987, n_controls_ =3206), Korean (n_cases_= 243, n_controls_=4041) and Thai (n_cases_=65, n_controls_=94) population were completed at The University of Tokyo, Japan. The Japanese and Korean GWAS data have been previously reported^22,23^. The Thai dataset was genotyped with the Axiom array.

### Dataset QC, imputation and GWAS

Quality control, imputation, and GWAS were conducted separately for each study location and population. GC lambda (GC_θ_) was used to assess inflation in all studies. The final case and control sizes and the number of variants tested can be found in Table S1 and Figure S1. Figure S11 shows matching of cases and controls in PCA plots. Manhattan plots and GC can be found in Figure S12 and genome-wide significant hits resulting from dataset GWAS are in table S13.

#### *GWAS data from NEPHROVIR/EU*: EU-European, EU-African, Maghrebian, Indian

Each file was quality controlled separately to remove related individuals (IBD > 0.1875), low call rate (genotype rate < 98%), and cases with discordant sex. SNPs were quality controlled for allele frequency (MAF < 0.01), call rate (genotype rate < 98%) in all cohorts, and Hardy Weinberg equilibrium (HWE P<1×10^−5^) in controls only. The EU-European datasets were generated in multiple files and were merged stepwise on the common subset of SNPs, with the previous QC procedure reapplied after each merge. PCA plots were constructed from PLINK v1.9 to identify population outliers and check for batch effects^58^. Pre-imputation QC was conducted using McCarthy Tools with the TOPMed reference panel to check strand alignment and allele assignment. Insertions and deletions were excluded prior to imputation. Each population was imputed separately and cases and controls were imputed together on the TOPMed Imputation Server with the TOPMed r2 reference panel^59–61^. The QC was repeated after imputation and SNPs with low imputation quality (rsq < 0.3) were excluded. After imputation, UCSC Liftover^62^ was used to convert SNP positions from each population dataset to build GRCh37 to match the build of summary statistics from other analyses. The association tests were completed using PLINK v1.9 under an additive model with principal component adjustment to account for population stratification.

#### GWAS data from Columbia University (US Cohorts)

US-European, US-African, US-South Asian, US-East Asian and Admixed American Population was assigned by KING^63^ kinship analysis software and based on continental population as defined by the 1000 Genomes Project for all cases and controls^64,65^. Within each continental population (EUR, AFR, AMR, SAS and EAS), we removed variants with genotype rate < 99%, MAF < 0.01 and HWE P<1×10^−5^. Each population was imputed separately with the TOPMed r2 panel^59–61^. After imputation, we removed first-degree relatives using KING, and variants with R2 < 0.8, MAF < 0.01 and HWE P<1×10^−5^. Principal components were calculated with FlashPCA^66^. For cohorts with large case/control imbalances (Admixed American and US-African), we used the SAIGE logistic model^65^ for calculating P-value and generating summary statistics. Association tests for European, South and East Asian were completed using PLINK v1.9 under an additive model with principal component adjustment to account for population stratification^58^.

#### *GWAS data from Kobe University*: Japanese, Korean and Thai

Quality control and analysis of the Japanese and Korean datasets are previously described in Jia et.al^23^. For the Thai analysis, SNPs were removed that had info score < 0.9, MAF < 0.005, call rate < 97%, or HWE P < 1×10^−5^. Individuals with missing rate > 3%, IBD > 0.1875 and PCA outliers were removed. Genotypes were imputed with The 1000 Genomes reference panel using SHAPEIT^67^ and IMPUTE2^68^. Logistic regression was performed with Plink1.9 and p-values were adjusted for genomic control (GC).

### Population-specific and multi-population meta-analysis

For each population-specific meta-analysis and the multi-population one, we conducted an inverse-variance, fixed-effect meta-analysis using METAL with adjustment for population stratification (GC) on each input dataset and assessment for heterogeneity selected^69^. For within-population meta-analyses, we removed variants with heterogeneity P-value < 0.05. All significant associations were visually inspected and single SNPs that did not follow the expected LD trend were not taken forward for further consideration.

For the European meta-analysis, we included available summary statistics of suggestive SNPs from Dufek et.al^24^, increasing the European sample size to 1,096 cases and 12,459 controls. The resulting suggestive and genome-wide significant SNPs are annotated in tables and figures.

### Multi-population meta-regression with MR-MEGA

To account for and assess heterogeneous loci, we conducted a meta-regression using MR-MEGA^27^. We included three principal components, which captured the population structure across all 12 datasets. This allowed us to stratify heterogeneity into residual heterogeneity and heterogeneity that correlates with population. We visualized each dataset’s PCs from MR-MEGA with the dataset-specific log odds ratio from METAL for each variant with heterogeneity that correlated with population. We adjusted for genomic control at the study level and after meta-regression to account for population structure within and between datasets. SNPs present in less than five studies were excluded. GC lambda (GC_θ_) was used to assess inflation. Results tables include summaries from both METAL and MR-MEGA analyses (Table 1). All Manhattan plots were generated with the qqman R package [doi: 10.21105/joss.00731.] and LocusZoom web tool^70^. All significant loci are > 1Mb from each other with r^2^ < 0.1. Loci were labeled by nearest genes.

### Conditional analyses

To identify independent secondary signals at the candidate loci, we used GCTA COJO^71,72^ to conduct approximate conditional analyses based on cohort-specific meta-analysis summary statistics. Conditional analysis was conducted in each dataset, with an LD reference generated from the dataset samples, due to differences in linkage disequilibrium structure between continental populations. Each cohort was conditioned for the eight independent loci identified from the initial meta-analysis. Multi-population meta-analysis of the conditioned cohorts was repeated in METAL^69^ to assess multi-population genome-wide significant secondary loci after GCTA.

### Heritability estimates

SNP-based heritability was estimated on a liability scale with LD score regression (LDSC)^73^ using a population prevalence of 16/100,000 and excluding HLA [chr6:25,000,000-34,000,000]. We used non-GC corrected population-specific meta-analysis summary statistics from METAL and pre-computed LD scores generated from 1000 Genomes EUR or EAS samples (https://alkesgroup.broadinstitute.org/LDSCORE/).

### Colocalization of SSNS GWAS variants and eQTLs datasets

We used fast enrichment estimation aided colocalization analysis (fastENLOC)^74^ for colocalization analysis with glomerular (n=240) and tubulointerstitial (n=311) eQTLs from nephrotic syndrome patients^55^, GTEx tissues (varied sample sizes), and immune eQTLs from both Blueprint^45^ (n=200) and DICE ^44^ (n=91) databases. Posterior probabilities for SSNS GWAS variants were calculated from MR-MEGA Z-scores using TORUS^75^. We used an LD panel from European and East Asian 1000 Genomes samples to define haplotype blocks in the pSSNS meta-analysis^56^. Enrichment of pSSNS GWAS variants in each eQTL dataset was estimated using fastENLOC and subsequently informed prior probabilities for each analysis. For colocalization with our kidney eQTLs, which had available raw data, we could identify multiple eQTL signals per gene and multiple colocalized signals at each locus. For all other data, in which only summary statistics were available, we assumed at most one colocalized signal at each locus and did not account for LD.

### Open chromatin annotation of credible sets

95% credible sets were constructed for each independent locus identified from the multi-population meta-regression from Bayes’ factors reported by MR-MEGA. Posterior inclusion probability (PIP) was estimated by dividing each Bayes’ factor by the summation of Bayes’ factors across all variants within a +/- 1Mb from the lead locus^76^.

SNPs within 95% credible sets of our genome-wide significant loci were evaluated for positional overlap based on the boundaries of known open chromatin peaks in kidney^49^ and immune^77^ cell types. Open chromatin peaks were identified by MACS2 peak calling algorithm and optimized by gkmQC^50^.

### HLA imputation and analysis

To fine-map the HLA region, we conducted HLA imputation with the four-digit multi-ethnic v2 reference panel on Michigan Imputation Server^51^. Cohorts were imputed individually to optimize population-specific structure within the HLA region. The imputed cohorts were then merged for multi-population associations. We used HLA-TAPAS ‘assoc’ module to conduct a logistic regression of the HLA region of the multi-population and population-specific datasets. For population-specific analyses, we adjusted for genotype-based principal components from Plinkv1.9^58^. The population-specific principal components and continental populations were included as covariates in the multi-population analysis. HLA-TAPAS was also used to conduct a stepwise conditional analysis, conditioning on the locus with the smallest association P-value. We additionally performed an omnibus test on the population-specific and multi-population cohorts to assess significance by amino acid position.

### HLA modeling

To predict the reference (with arginine at position 47 and serine at position 52) structure of *HLA-DQA1* we extracted the sequence of *HLA-DQA1* from UNIPROT database (Uniprot ID: P01909). We used NCBI BLASTp against PDB database to find the closest structure associated with the amino acid sequence of P01909. We identified the top hit as 6PX6_A (HLA-TCR complex, E = 2e-161) for the *HLA-DQA1* sequence^78^. We extracted the PDB coordinates for chain A from the 6PX6 and visualized in PYMOL v2.5. Since the most common amino acid haplotype in the control population was arginine (47) and serine (52), we performed mutagenesis using PYMOL to model the reference protein 3-D structure^79^.

In brief, we used the mutagenesis tool from PYMOL and selected the rotamer (most likely amino acid conformation) for arginine and serine which showed the minimum number of clashes with nearby atoms. Afterwards, we adjusted the conformation of nearby atoms (within 5 Angstrom) to minimum free state using “Clean” command in PyMOL which uses MMFF94 force field^80^. Though point mutations locally affect the conformation of the protein, they can result in torsion, bending and stretching of the entire molecule. Therefore, we exported the protein structure to SPDBV software for further refinement^81^.

We first fixed all the side chains of all amino acids to the best rotamer conformation using the simulated annealing method. Afterwards, we performed energy minimization using GROMOS 96 force field to extract the 3D coordinates that represent the lowest minimum energy conformation^82^. The refined protein structure of *HLA-DQA1* was then assessed for changes in stability of protein for both amino acid combinations for each haplotype using “MULTIPLE MUTATION” in DynMut2 server^54^. The instability of HLA-DQA1 was evaluated using the predicted ddG parameter which measures changes in Gibbs free energy between the folded and unfolded states and the change in folding when a mutation is present. The interaction among amino acids in reference and mutated structure were predicted using Arpeggio^83^ and visualized in PyMOL.

### Polygenic risk score analysis

#### Construction of the polygenic risk score (PRS)

To investigate genetic risk across the genome, we generated a polygenic risk score (PRS) from the GWAS of European (US-European) and East Asian (Japanese, Korean, US-East Asian) populations using PRS-CSx^84^. Population-specific weights estimated by PRS-CSx were used as input, along with a testing/training dataset, for optimization of the gamma-gamma priors a and b and the global shrinkage parameter used in the PRS-CSx model. Our main objective was to test clinical associations with PRS within case cohorts. We used case/control data and prediction accuracy to choose the best PRS model parameters. We fine-tuned and estimated PRS population weights in the EU-European and NEPTUNE separately. Since NEPTUNE is a case-only cohort, we included population--matched controls from the 1000 Genomes Project. In each dataset, 80% of cases and controls were randomly selected for training and 20% for testing. We varied the PRS-CSx hyper-parameters and chose the combination that gave the best prediction accuracy. The prediction accuracy with an area under the receiver operating characteristic curve was 0.74 (95%CI: 0.72-0.77) in EU-European dataset and 0.64 (0.61-0.68) in NEPTUNE.

#### Clinical associations with PRS

The PRS was applied to pediatric participants from NEPTUNE^55^ (n=165), and the EU-European data for which clinical data were available (n=239). For bivariate tests, we used the Wilcoxon test for binary traits, Kruskal-Wallis for categorical traits and univariate linear regression for continuous traits. Population in NEPTUNE was predicted using Peddy^85^ and the 1000 Genomes reference panel. For associations with the age of onset, we conducted linear regression adjusting for sex, four genetic principal components, and histology (NEPTUNE only).

## Data Availability

All data produced in this present study are available upon reasonable request to the authors and will be subsequently uploaded to a data-sharing portal.

