## Supplement for "Multi-population genome-wide association study implicates both immune and non-immune factors in the etiology of pediatric steroid sensitive nephrotic syndrome"

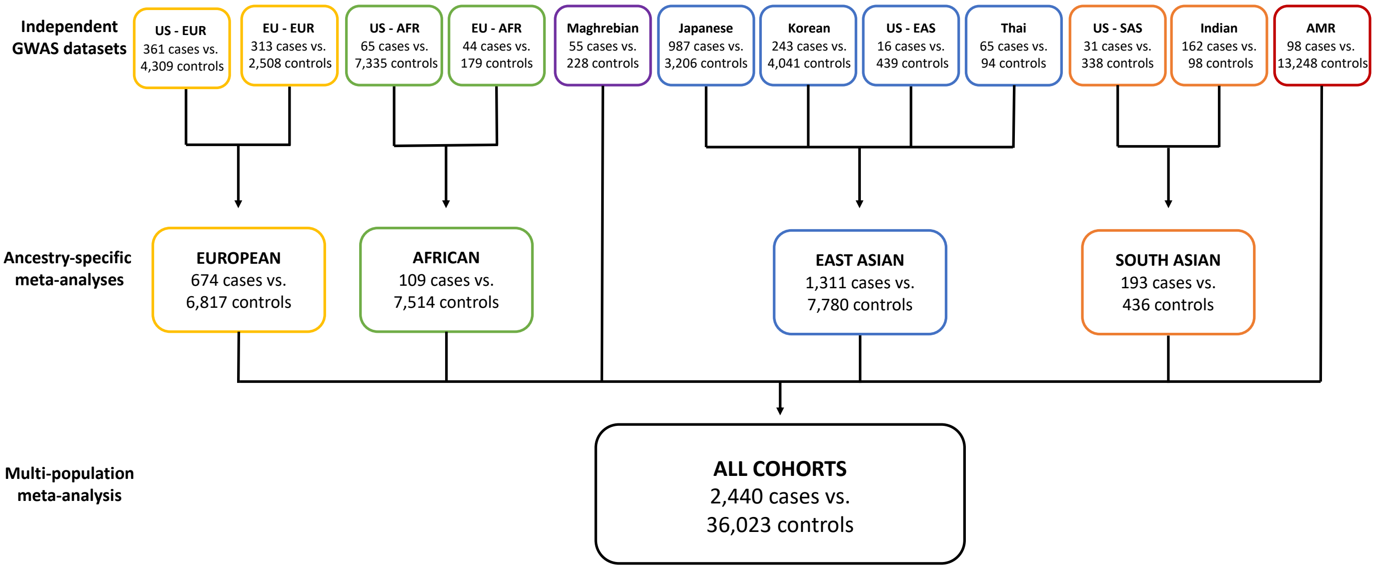


**Figure S1. Flow-chart of GWAS.** EUR= European, AFR= African, EAS= East Asian, SAS = South Asian

**Table S1.** Summary of GWAS datasets

| GWAS Cohort | Ancestry | n cases | n controls | n total | GC-lambda | PCs | n SNPs |
|---|---|---|---|---|---|---|---|
| Nephrovir/EU | European | 313 | 2,508 | 2,821 | 1.05 | 3 | 8,112,877 |
| US Cohort | European | 361 | 4,309 | 4,670 | 1.06 | 5 | 8,316,416 |
| US Cohort | African | 65 | 7,335 | 7,400 | 0.84 | 2 | 13,421,506 |
| Nephrovir/EU | African | 44 | 179 | 223 | 1.02 | 5 | 12,413,167 |
| US Cohort | East Asian | 16 | 439 | 455 | 1.00 | 4 | 6,196,585 |
| Japanese | East Asian | 987 | 3,206 | 4,193 | 1.05 | 4 | 6,088,373 |
| Korean | East Asian | 243 | 4,041 | 4,284 | 1.03 | 0 | 2,912,065 |
| Thai | East Asian | 65 | 94 | 159 | 1.00 | 0 | 4,946,221 |
| US Cohort | South Asian | 31 | 338 | 369 | 0.93 | 3 | 7,637,355 |
| Indian | South Asian | 162 | 98 | 260 | 1.03 | 3 | 8,159,073 |
| Maghrebian | Maghrebian | 55 | 228 | 283 | 0.90 | 3 | 10,072,905 |
| Admixed American | Admixed American | 98 | 13,248 | 13,346 | 0.98 | 3 | 9,264,840 |
| Total | | 2,440 | 36,023 | 38,463 | | | |

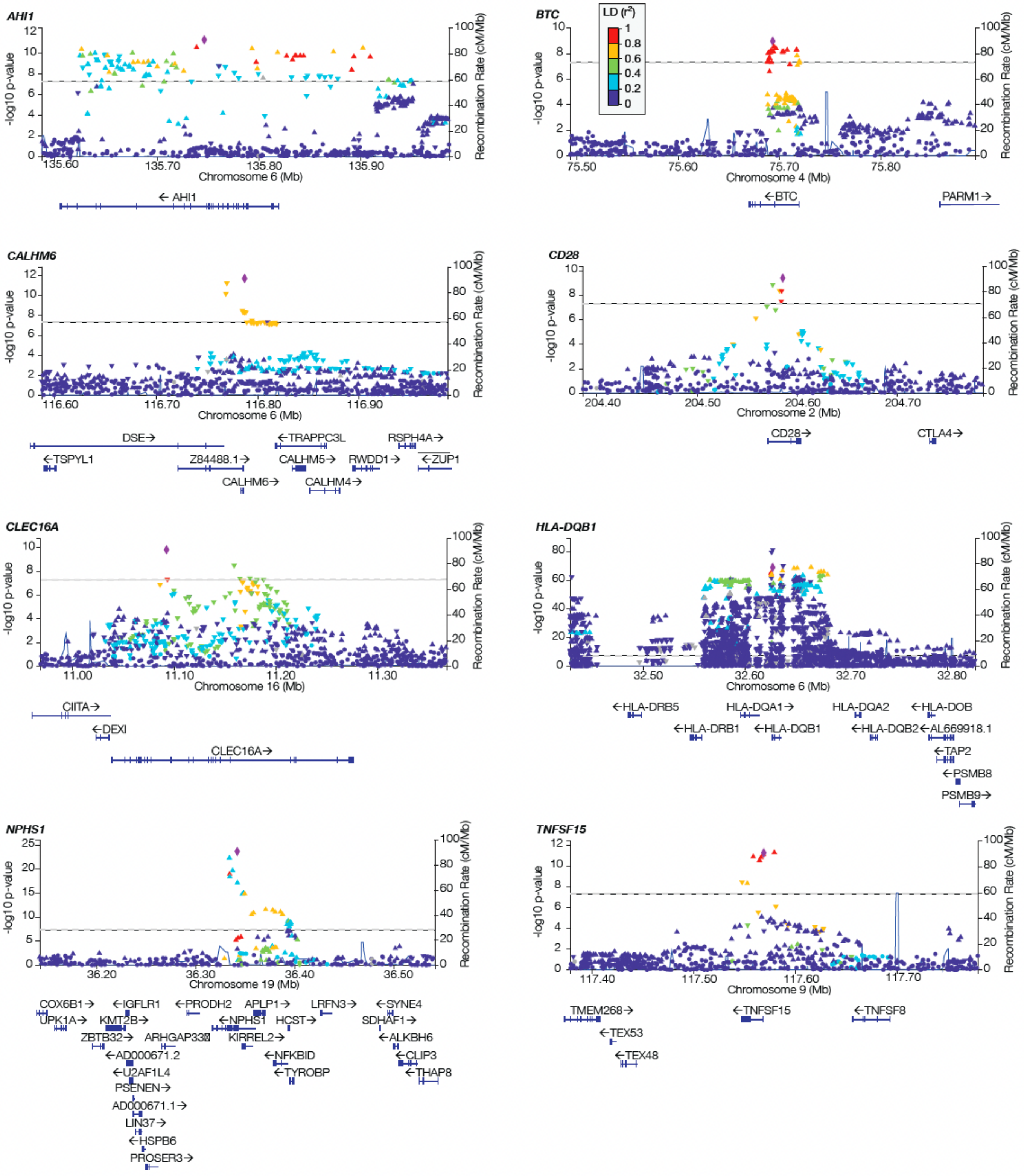


**Figure S2: 'LocusZoom' of genome-wide significant loci.**

Each figure is labeled by closest gene. Linkage disequilibrium (LD/$r^2$) with the most significant SNP in the reference panel (purple diamond) is estimated from all 1000 genomes populations. P-values are from MR-MEGA association. Horizontal line indicates genome-wide significance threshold ($5x10^{-8}$).

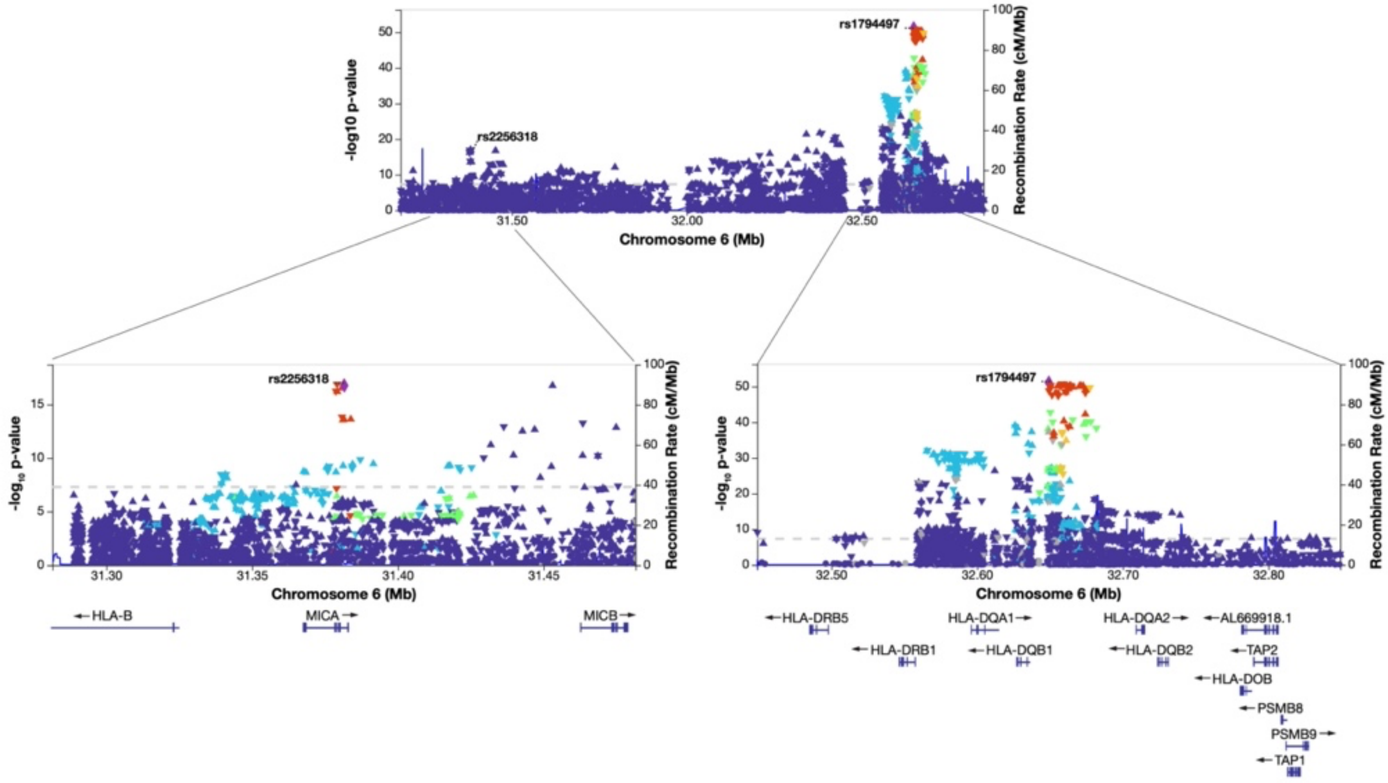


**Figure S3. 'LocusZoom' of significant loci from conditional analysis.** Conditioned on: rs55730955, rs28862935 ,rs1063355 , rs2637681, rs7759971, rs10817678 , rs8062322, rs56117924. rs2256318 and rs1794497 are ~1.3Mb apart with $r^2$=0.04 in all 1,000 Genomes samples.

**Table S2. Summary of population-specific meta-analyses.**

| GWAS Cohort | n cases | n controls | n total | GC-lambda | n SNPs |
|---|---|---|---|---|---|
| African | 109 | 7,514 | 7,623 | 0.85 | 14,302,064 |
| East Asian | 1,311 | 7,780 | 9,091 | 0.99 | 8,644,038 |
| European | 674 | 6,817 | 7,491 | 1.07 | 8,414,111 |
| South Asian | 193 | 436 | 629 | 0.95 | 8,460,574 |

**Table S3: Genome-wide significant SNPs from population-specific meta-analyses**

All SNP are > 1Mb from each other with $r^2 < 0.1$. Variants showing high within population heterogeneity are removed (HetPVal < 0.05). We found no genome-wide significant associations in the South Asian, African, Admixed American and Maghrebian meta-analyses.

| Population | Top SNP | Position (hg19) | Nearest Gene | EA | NEA | OR [95% CI] | P |
|---|---|---|---|---|---|---|---|
| East Asian | rs9274740 | 6:32637968 | *HLA-DQB1* | A | T | 0.41 [0.36, 0.47] | $1.25 \times 10^{-39}$ |
| East Asian | rs412175 | 19:36342103 | *NPHS1* | T | C | 0.53 [0.47, 0.60] | $7.47 \times 10^{-25}$ |
| East Asian | rs2596485 | 6:31364870 | *MICA* | T | C | 0.61 [0.53, 0.69] | $2.46 \times 10^{-15}$ |
| East Asian | rs7848647 | 9:117569046 | *TNFSF15* | T | C | 0.70 [0.63, 0.77] | $9.57 \times 10^{-13}$ |
| East Asian | rs1181388 | 2:204575951 | *CD28* | A | G | 0.73 [0.66, 0.80] | $4.93 \times 10^{-11}$ |
| East Asian | rs115180879 | 6:28984623 | *ZNF311* | T | C | 3.00 [2.09, 4.32] | $3.10 \times 10^{-9}$ |
| East Asian | rs8062322 | 16:11092319 | *CLEC16A* | A | C | 0.68 [0.59, 0.77] | $5.48 \times 10^{-9}$ |
| European | rs9271747 | 6:32626037 | *HLA-DQB1* | T | G | 3.30 [2.82, 3.86] | $5.69 \times 10^{-51}$ |
| European | rs2637678* | 6:116787378 | *CALHM6* | T | C | 1.74 [1.58, 1.93] | $2.35 \times 10^{-27}$ |
| European | rs2857607 | 6:31517248 | *NFKBIL1* | T | C | 1.95 [1.66, 2.31] | $1.18 \times 10^{-13}$ |
| European | rs2746419* | 6:135653855 | *AHI1* | A | C | 1.34 [1.22, 1.47] | $1.63 \times 10^{-10}$ |
| European | rs111796602* | 7:37402490 | *ELMO1* | T | C | 0.68 [0.59, 0.78] | $1.72 \times 10^{-8}$ |
| European | rs12911841 | 15:79162355 | *MORF4L1* | T | C | 2.95 [2.01, 4.34] | $3.88 \times 10^{-8}$ |

*European meta-analysis includes summary statistics from the GWAS Catalogue from Dufek et.al., for a total of 1,096 cases and 12,459 controls.

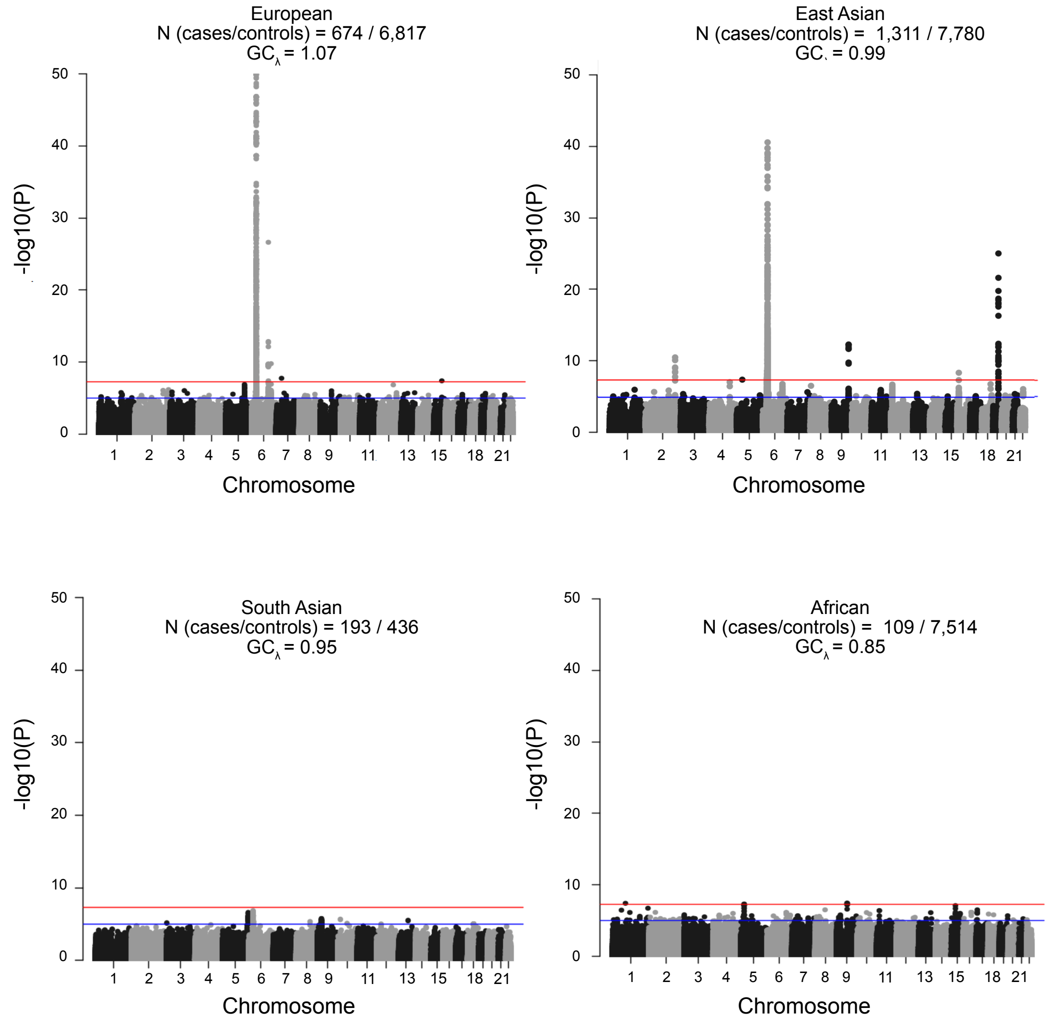


**Figure S4. Manhattan plots of ancestry-specific meta-analyses.**
Red line indicated genome-wide significance threshold ($5 \times 10^{-8}$); blue line is suggestive significance ($5 \times 10^{-5}$). Number of cases and controls and genomic controls lambda are reported for each dataset.

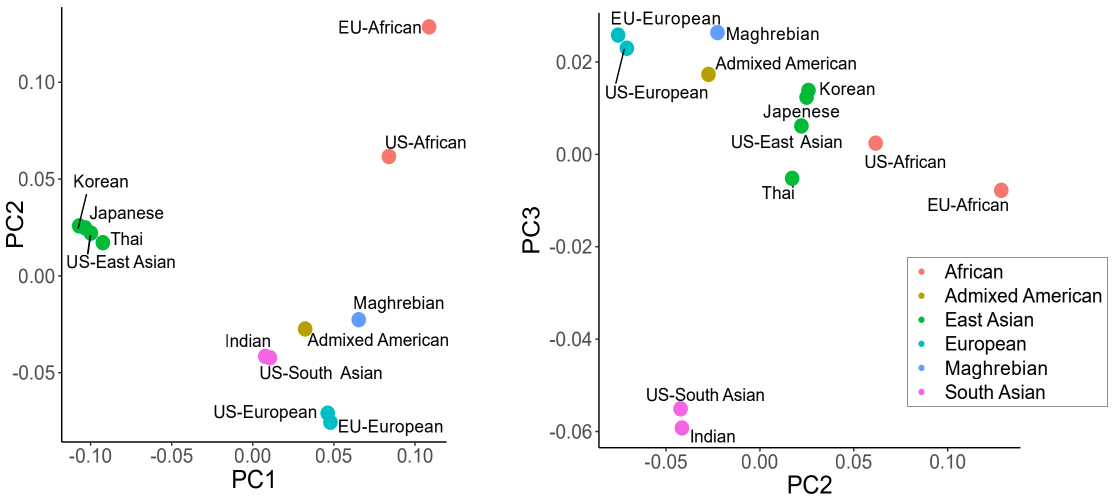


**Figure S5. Dataset principal components (PCs) used in MR-MEGA meta-regression analysis.** Labeled by dataset, colored by population. PCs based off allele frequencies across all GWAS variants.

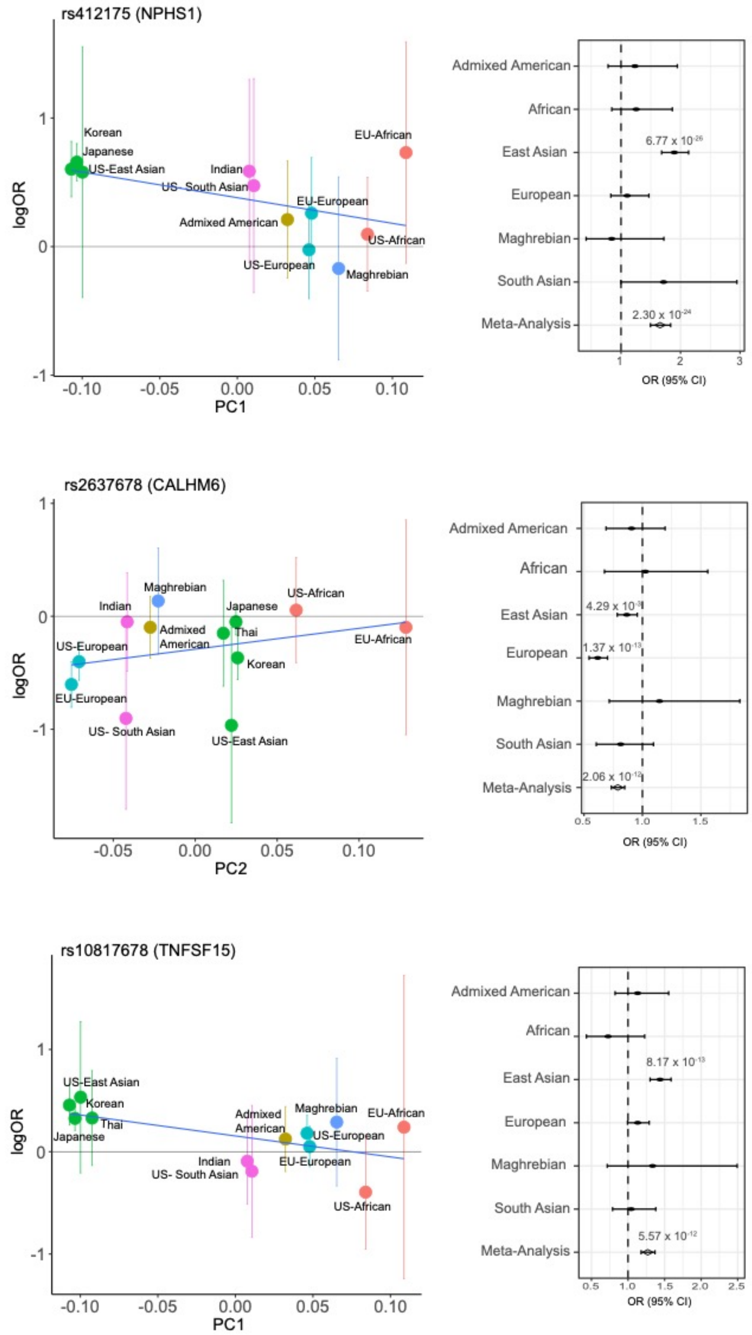


**Figure S6. Population-specificity of heterogeneous GWAS variants**. Left: correlation of MR-MEGA PCs and logOR for each dataset. Each SNP is labeled with the nearest gene. Right: Forrest plot for population analysis. Significant odds ratios are labeled. logOR = logarithm of the odds ratio, OR [95%CI] = odds ratio with 95% confidence interval.

**Table S4. Suggestive Significant SNPs, Multi-population Meta-analysis.**

| | | | | | Fixed-effects meta-analysis | | | Meta-regression (MR-MEGA) | |
|---|---|---|---|---|---|---|---|---|---|
| Nearest Gene* | Top SNP | Position (hg19) | EA | NEA | OR [95% CI] | P-value | Het $I^2$ P-value | P-value | Ancestry het P-value |
| *GSDMB* | rs9303279 | 17:38073968 | G | C | 1.22 [1.13, 1.31] | $1.88 \times 10^{-7}$ | 0.16 | $2.49 \times 10^{-7}$ | 0.05 |
| *TNSFS4* | rs1012507 | 1:173219471 | T | G | 1.20 [1.11, 1.29] | $5.73 \times 10^{-6}$ | 0.05 | $2.92 \times 10^{-7}$ | $2.52 \times 10^{-3}$ |
| *DHX15** | rs16875349 | 4:24215917 | C | T | 1.34 [1.21, 1.48] | $1.09 \times 10^{-8}$ | 0.01 | $3.56 \times 10^{-7}$ | 0.81 |
| *PHC1* | rs1805732 | 12:9090892 | T | C | 0.82 [0.76, 0.88] | $1.71 \times 10^{-7}$ | 0.51 | $4.65 \times 10^{-7}$ | 0.10 |
| *MORF4L1* | rs12911841 | 15:79162355 | T | C | 1.82 [1.43, 2.30] | $6.92 \times 10^{-7}$ | 0.03 | $5.82 \times 10^{-7}$ | 0.03 |
| *CLIC4* | rs4649032 | 1:25197586 | T | C | 1.20 [1.12, 1.30] | $1.18 \times 10^{-6}$ | 0.16 | $1.18 \times 10^{-6}$ | 0.03 |
| *KC6* | rs16975006 | 18:39160883 | T | G | 1.68 [1.39, 2.02] | $6.21 \times 10^{-8}$ | 0.39 | $1.64 \times 10^{-6}$ | 0.38 |
| *C16orf95* | rs7197567 | 16:86980044 | T | C | 1.31 [1.18, 1.45] | $3.85 \times 10^{-7}$ | 0.75 | $2.69 \times 10^{-6}$ | 0.23 |
| *COCH* | rs12431424 | 14:31318179 | A | T | 0.78 [0.7, 0.87] | $3.67 \times 10^{-6}$ | 0.15 | $3.14 \times 10^{-6}$ | 0.03 |
| *SRBD1** | rs78341222 | 2:45539561 | A | G | 1.51 [1.15, 1.99] | $3.27 \times 10^{-3}$ | $5.48 \times 10^{-4}$ | $3.71 \times 10^{-6}$ | $6.09 \times 10^{-5}$ |
| *BAD* | rs477895 | 11:64048912 | C | T | 1.19 [1.1, 1.29] | $3.44 \times 10^{-5}$ | 0.07 | $3.81 \times 10^{-6}$ | $4.56 \times 10^{-3}$ |
| *RRP12* | rs6584128 | 10:99171926 | G | T | 1.21 [1.13, 1.3] | $1.74 \times 10^{-7}$ | 0.19 | $5.10 \times 10^{-6}$ | 0.75 |
| *NFIA* | rs75447844 | 1:61807924 | T | C | 1.71 [1.3, 2.24] | $1.12 \times 10^{-4}$ | $4.69 \times 10^{-3}$ | $7.36 \times 10^{-6}$ | $2.58 \times 10^{-3}$ |

* Closest gene was not protein coding. SNPs with P-value > 0.05 in METAL were excluded.

**Table S5. Allele frequency of genome-wide and suggestive significant SNPs across ancestries.**

| | | | European | | South Asian | | East Asian | | African | | Maghrebian | | Admixed American | |
|---|---|---|---|---|---|---|---|---|---|---|---|---|---|---|
| Nearest Gene | Top SNP | EA | Case | Control | Case | Control | Case | Control | Case | Control | Case | Control | Case | Control |
| *HLA-DQB1* | rs1063355 | T | 0.22 | **0.40** | 0.35 | **0.49** | 0.28 | **0.44** | 0.28 | **0.46** | 0.15 | **0.38** | 0.17 | **0.34** |
| *NPHS1* | rs412175 | C | **0.05** | 0.04 | **0.10** | 0.06 | **0.24** | 0.15 | **0.46** | 0.34 | 0.12 | **0.14** | **0.18** | 0.15 |
| *CALHM6* | rs2637678 | C | 0.33 | **0.43** | 0.21 | **0.26** | 0.40 | **0.44** | 0.22 | **0.29** | **0.52** | 0.50 | **0.43** | 0.38 |
| *AHI1* | rs7759971 | T | **0.40** | 0.35 | **0.42** | 0.39 | **0.26** | 0.22 | 0.19 | **0.21** | **0.35** | 0.21 | **0.37** | 0.30 |
| *TNSFS15* | rs10817678 | G | 0.29 | **0.33** | **0.26** | 0.22 | 0.34 | **0.44** | 0.13 | **0.17** | 0.15 | **0.18** | 0.23 | **0.24** |
| *CLEC16A* | rs8062322 | A | 0.31 | **0.34** | 0.30 | **0.38** | 0.14 | **0.19** | **0.46** | 0.44 | 0.20 | **0.34** | **0.33** | 0.30 |
| *CD28* | rs55730955 | A | 0.05 | **0.06** | 0.09 | 0.09 | 0.45 | **0.55** | 0.07 | **0.08** | 0.05 | **0.10** | 0.08 | **0.11** |
| *BTC* | rs28862935 | A | **0.23** | 0.16 | **0.28** | 0.25 | **0.06** | 0.05 | **0.48** | 0.36 | **0.44** | 0.30 | **0.34** | 0.23 |
| *GSDMB* | rs9303279 | G | **0.50** | 0.45 | 0.41 | **0.42** | **0.30** | 0.26 | 0.11 | **0.21** | **0.42** | 0.33 | **0.35** | 0.34 |
| *TNSFS4** | rs1012507 | T | **0.37** | 0.34 | 0.32 | 0.32 | **0.25** | 0.23 | 0.24 | **0.29** | 0.21 | **0.26** | 0.34 | **0.39** |
| *DHX15** | rs16875349 | C | **0.07** | 0.06 | **0.14** | 0.12 | **0.24** | 0.18 | **0.17** | 0.14 | 0.05 | **0.06** | 0.06 | **0.09** |
| *PHC1* | rs1805732 | T | 0.40 | **0.43** | 0.47 | 0.47 | 0.42 | **0.48** | 0.15 | **0.21** | 0.30 | **0.38** | 0.35 | **0.38** |
| *MORF4L1* | rs12911841 | T | **0.03** | 0.01 | - | - | - | - | **0.27** | 0.18 | 0.18 | 0.18 | **0.10** | 0.05 |
| *CLIC4* | rs4649032 | T | **0.50** | 0.49 | **0.43** | 0.35 | **0.49** | 0.44 | **0.44** | 0.40 | 0.47 | **0.48** | **0.44** | 0.41 |
| *KC6* | rs16975006 | T | **0.03** | 0.02 | **0.04** | 0.02 | 0.01 | 0.01 | **0.25** | 0.20 | **0.12** | 0.06 | **0.12** | 0.06 |
| *C16orf95* | rs7197567 | T | **0.09** | 0.06 | 0.06 | **0.07** | **0.21** | 0.18 | **0.54** | 0.42 | **0.23** | 0.19 | **0.22** | 0.20 |
| *COCH* | rs12431424 | A | 0.13 | **0.15** | 0.10 | **0.15** | 0.16 | **0.18** | 0.12 | **0.19** | **0.12** | 0.10 | 0.09 | **0.21** |
| *SRBD1** | rs78341222 | A | 0.03 | 0.03 | **0.05** | 0.04 | - | - | - | - | **0.07** | 0.03 | **0.05** | 0.02 |
| *BAD* | rs477895 | C | **0.19** | 0.18 | 0.26 | **0.27** | **0.25** | 0.21 | **0.56** | 0.43 | **0.49** | 0.34 | **0.31** | 0.21 |
| *RRP12* | rs6584128 | G | **0.37** | 0.34 | **0.42** | 0.40 | **0.36** | 0.30 | **0.64** | 0.53 | **0.55** | 0.51 | **0.43** | 0.37 |
| *NFIA* | rs75447844 | T | 0.02 | 0.02 | **0.09** | 0.05 | 0.13 | **0.15** | - | - | 0.01 | 0.01 | 0.06 | **0.14** |

* Closest gene was not protein coding. The group with higher frequency is indicated in bold. EA = effect allele

**Table S6. SNPs that significantly colocalize with tissue and cell type eQTLs.**
Top colocalized SSNS GWAS / eQTL loci (RCP > 0.2). Each row represents a SSNS associated SNP that also associates with tissue/cell-type gene expression. The top SNP (highest SCP) for each gene-tissue association is included. RCP = regional colocalization probability, SCP = SNP colocalization probability.

| RS ID | Position (hg19) | Ensembl Gene ID | Gene Symbol | Study | Tissue/Cell type | RCP | SCP |
|---|---|---|---|---|---|---|---|
| rs7759971 | 6:135746884 | ENSG00000135541 | *AHI1* | DICE | Monocytes, classical | 0.65 | 0.23 |
| rs6908428 | 6:135793706 | ENSG00000135541 | *AHI1* | DICE | T cell, CD4, memory TREG | 0.65 | 0.62 |
| rs6908428 | 6:135793706 | ENSG00000135541 | *AHI1* | DICE | T cell, CD4, naive | 0.62 | 0.62 |
| rs6908428 | 6:135793706 | ENSG00000135541 | *AHI1* | DICE | T cell, CD4, naive TREG | 0.57 | 0.57 |
| rs6908428 | 6:135793706 | ENSG00000135541 | *AHI1* | DICE | T cell, CD4, TFH | 0.56 | 0.56 |
| rs6908428 | 6:135793706 | ENSG00000135541 | *AHI1* | DICE | B cell, naive | 0.56 | 0.56 |
| rs6908428 | 6:135793706 | ENSG00000135541 | *AHI1* | DICE | Monocytes, non-classical | 0.53 | 0.52 |
| rs6908428 | 6:135793706 | ENSG00000135541 | *AHI1* | DICE | T cell, CD4, TH17 | 0.51 | 0.51 |
| rs6908428 | 6:135793706 | ENSG00000135541 | *AHI1* | DICE | T cell, CD8, naive | 0.40 | 0.40 |
| rs6908428 | 6:135793706 | ENSG00000135541 | *AHI1* | DICE | T cell, CD4, TH1/17 | 0.39 | 0.39 |
| rs7759971 | 6:135746884 | ENSG00000135541 | *AHI1* | BluePrint | Monocyte | 0.26 | 0.10 |
| rs2637678 | 6:116787378 | ENSG00000188820 | *CALHM6* | DICE | Monocytes, classical | 0.86 | 0.86 |
| rs2637681 | 6:116769845 | ENSG00000188820 | *CALHM6* | GTEx | Adipose visceral omentum | 0.39 | 0.39 |
| rs2637681 | 6:116769845 | ENSG00000188820 | *CALHM6* | GTEx | Skin (not sun exposed) | 0.25 | 0.25 |
| rs8076131 | 17:38080912 | ENSG00000073605 | *GSDMB* | DICE | T cell, CD4, memory TREG | 0.29 | 0.16 |
| rs5024432 | 6:32684468 | ENSG00000237541 | *HLA-DQA1* | GTEx | Skin (not sun exposed) | 0.22 | 0.13 |
| rs6908428 | 6:135793706 | ENSG00000231028 | *LINC00271* | DICE | T cell, CD4, memory TREG | 0.36 | 0.07 |
| rs6908428 | 6:135793706 | ENSG00000234084 | *Lnc-MYB-2* | DICE | T cell, CD4, naive | 0.66 | 0.26 |
| rs6908428 | 6:135793706 | ENSG00000234084 | *Lnc-MYB-3* | DICE | B cell, naive | 0.52 | 0.51 |
| rs7759971 | 6:135746884 | ENSG00000234084 | *Lnc-MYB-4* | DICE | Monocytes, non-classical | 0.42 | 0.17 |
| rs6908428 | 6:135793706 | ENSG00000234084 | *Lnc-MYB-5* | DICE | T cell, CD4, TH1/17 | 0.33 | 0.32 |
| rs6908428 | 6:135793706 | ENSG00000234084 | *Lnc-MYB-6* | DICE | T cell, CD4, TFH | 0.33 | 0.20 |
| rs6908428 | 6:135793706 | ENSG00000234084 | *Lnc-MYB-7* | DICE | T cell, CD4, naive TREG | 0.31 | 0.19 |
| rs6908428 | 6:135793706 | ENSG00000234084 | *Lnc-MYB-8* | DICE | T cell, CD4, memory TREG | 0.22 | 0.09 |
| rs6908428 | 6:135793706 | ENSG00000234084 | *Lnc-MYB-9* | DICE | T cell, CD4, TH17 | 0.21 | 0.10 |
| rs12941333 | 17:38040534 | ENSG00000172057 | *ORMDL3* | DICE | T cell, CD4, memory TREG | 0.23 | 0.02 |
| rs7848647 | 9:117569046 | ENSG00000181634 | *TNFSF15* | DICE | Monocytes, classical | 0.95 | 0.28 |
| rs10817678 | 9:117579457 | ENSG00000181634 | *TNFSF15* | BluePrint | Monocyte | 0.90 | 0.69 |
| rs6478108 | 9:117558703 | ENSG00000181634 | *TNFSF15* | GTEx | Whole blood | 0.62 | 0.43 |
| rs6478109 | 9:117568766 | ENSG00000181634 | *TNFSF15* | GTEx | Artery_Aorta | 0.57 | 0.30 |
| rs7848647 | 9:117569046 | ENSG00000181634 | *TNFSF15* | DICE | Monocytes, non-classical | 0.22 | 0.05 |
| rs111922894 | 7:129178359 | ENSG00000158457 | *TSPAN33* | DICE | T cell, CD4, naive | 0.25 | 0.25 |

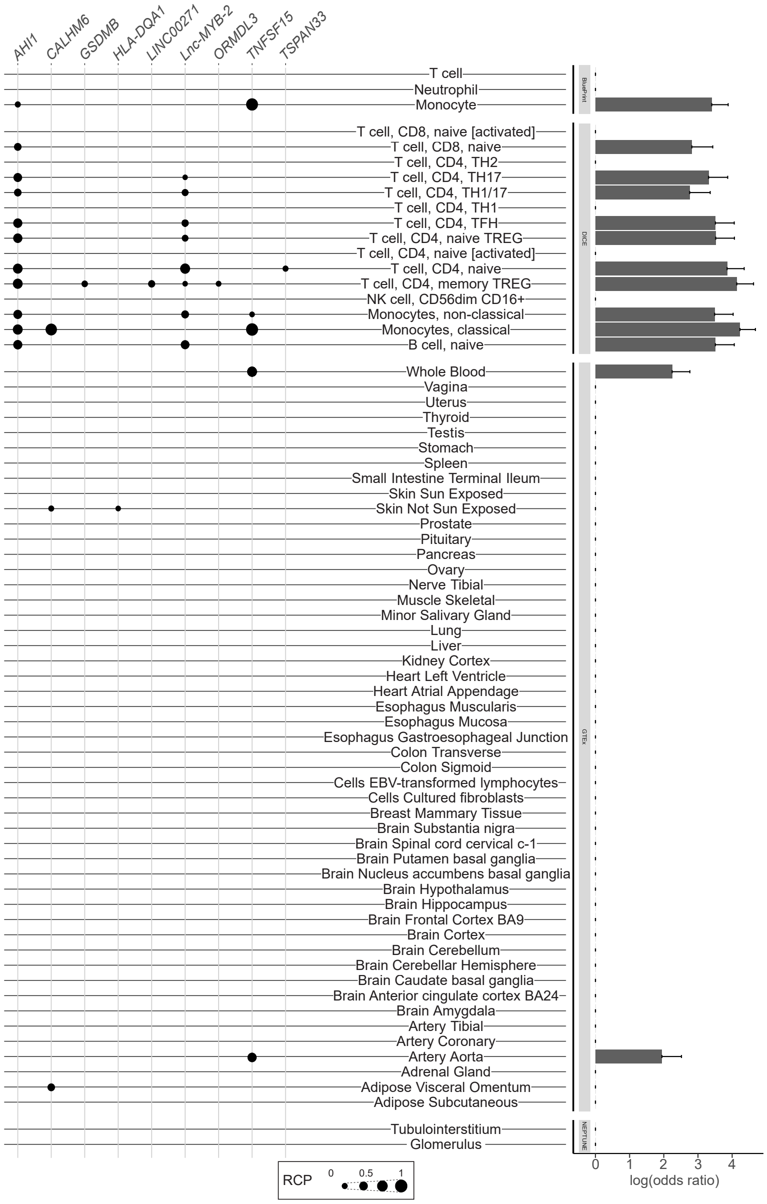


**Figure S7. Colocalization of SSNS GWAS and all eQTL datasets.** Each eQTL data set is labeled with colocalized loci to the right and enrichment estimates to the left. Genes with regional colocalization probability (RCP) > 0.2 in at least one tissue/cell are included. SSNS GWAS loci that colocalized with tissue/cell-type eQTLs are indicated by black dots, with larger dots indicating higher regional colocalization probability (RCP). Enrichment estimates, from fastENLOC, are based of genome-wide summary statistics from GWAS and include a shrinkage parameter, resulting in 0 enrichment for multiple tissues/cell-types.

**Table S7. Credible sets for genome-wide significant loci in multi-population meta-analysis with open chromatin annotation.**

| RS ID | Position (hg19) | EA | NEA | P-value (MR-MEGA) | PIP | Location | Closest Gene | Kidney open chromatin | Immune open chromatin |
|---|---|---|---|---|---|---|---|---|---|
| rs8062322 | 16:11092319 | A | C | $8.98 \times 10^{-12}$ | 0.93 | intronic | *CLEC16A* | | |
| rs9652601 | 16:11174365 | A | G | $1.18 \times 10^{-9}$ | 0.01 | intronic | *CLEC16A* | *POD,ENDO,PT1,PT2, PT3,PT, LH,PC,LEUK* | *CMP* |
| rs7204643 | 16:11167603 | C | T | $1.46 \times 10^{-9}$ | 0.01 | intronic | *CLEC16A* | *ENDO,PT1,PT2,PT,LH,DCT,CNT ,PC,ICB* | *GMP,LMPP,MEP* |
| rs887864 | 16:11158885 | G | A | $1.45 \times 10^{-9}$ | 0.01 | intronic | *CLEC16A* | | |
| rs7200940 | 16:11164567 | G | C | $1.56 \times 10^{-9}$ | 0.01 | intronic | *CLEC16A* | | |
| rs412175 | 19:36342103 | C | T | $2.29 \times 10^{-24}$ | 0.94 | Intronic | *NPHS1* | *ENDO, PT2* | |
| rs56117924 | 19:36334182 | A | G | $4.98 \times 10^{-23}$ | 0.05 | intronic | *NPHS1* | | |
| rs55730955 | 2:204585956 | A | T | $1.00 \times 10^{-11}$ | 0.72 | intronic | *CD28* | | *CD4,CD8* |
| rs4673259 | 2:204582623 | C | T | $3.95 \times 10^{-11}$ | 0.19 | intronic | *CD28* | | *CD4,CD34,CMP,GMP, HSC,LMPP,MEP* |
| rs3769684 | 2:204584759 | C | T | $1.22 \times 10^{-10}$ | 0.07 | intronic | *CD28* | *LEUK* | *CD4,CD8,NK* |
| rs28862935 | 4:75693465 | A | G | $4.29 \times 10^{-11}$ | 0.21 | intronic | *BTC* | | |
| rs10005089 | 4:75694606 | T | C | $9.38 \times 10^{-11}$ | 0.10 | intronic | *BTC* | *PEC,ENDO,PT1,PT2,PT3,PT,LH, DCT,CNT,PC,ICA,ICB,* | *HSC,LMPP,MEP,MPP* |
| rs6532431 | 4:75696612 | G | T | $1.50 \times 10^{-10}$ | 0.06 | intronic | *BTC* | | |
| rs28420261 | 4:75693713 | A | C | $1.77 \times 10^{-10}$ | 0.05 | intronic | *BTC* | | |
| rs4859427 | 4:75691379 | A | G | $1.92 \times 10^{-10}$ | 0.05 | intronic | *BTC* | | |
| rs60799154 | 4:75692264 | T | C | $2.05 \times 10^{-10}$ | 0.05 | intronic | *BTC* | | |
| rs28773846 | 4:75692704 | G | T | $2.20 \times 10^{-10}$ | 0.05 | intronic | *BTC* | | |
| rs72867560 | 4:75691141 | A | G | $2.49 \times 10^{-10}$ | 0.04 | intronic | *BTC* | | *ERY* |
| rs59882675 | 4:75689112 | G | A | $2.64 \times 10^{-10}$ | 0.04 | intronic | *BTC* | | |
| rs28478898 | 4:75689280 | G | C | $2.66 \times 10^{-10}$ | 0.04 | intronic | *BTC* | | |
| rs28472417 | 4:75689353 | C | T | $2.66 \times 10^{-10}$ | 0.04 | intronic | *BTC* | | |
| rs10009801 | 4:75694208 | C | T | $2.82 \times 10^{-10}$ | 0.03 | intronic | *BTC* | | |
| rs28577058 | 4:75716947 | A | G | $3.02 \times 10^{-10}$ | 0.03 | intronic | *BTC* | *POD,MESFIB,CNT,PC,ICA,ICB* | |
| rs28681122 | 4:75717024 | A | G | $3.38 \times 10^{-10}$ | 0.03 | intronic | *BTC* | | |
| rs10023748 | 4:75703330 | G | T | $3.89 \times 10^{-10}$ | 0.02 | intronic | *BTC* | | |
| rs28576102 | 4:75704143 | G | A | $4.18 \times 10^{-10}$ | 0.02 | intronic | *BTC* | | |
| rs72867585 | 4:75702876 | G | A | $4.30 \times 10^{-10}$ | 0.02 | intronic | *BTC* | | |
| rs28522727 | 4:75707658 | C | T | $4.33 \times 10^{-10}$ | 0.02 | intronic | *BTC* | | |
| rs6848247 | 4:75690027 | G | T | $9.82 \times 10^{-10}$ | 0.01 | intronic | *BTC* | | |
| rs28458910 | 4:75688543 | G | A | $9.97 \times 10^{-10}$ | 0.01 | intronic | *BTC* | | |
| rs28822209 | 4:75690673 | A | G | $1.09 \times 10^{-9}$ | 0.01 | intronic | *BTC* | | |
| rs28565087 | 4:75688563 | G | A | $1.10 \times 10^{-9}$ | 0.01 | intronic | *BTC* | | |
| rs2637681 | 6:116769845 | G | T | $3.27 \times 10^{-11}$ | 0.52 | intergenic | *CALHM6* | | |
| rs2637678 | 6:116787378 | C | T | $5.54 \times 10^{-11}$ | 0.31 | intergenic | *CALHM6* | | |
| rs2858829 | 6:116768917 | G | A | $6.27 \times 10^{-10}$ | 0.03 | intergenic | *CALHM6* | *LEUK* | *CD34,CMP,GMP,HSC,LMPP, MEP, MONO,MPP* |
| rs61449271 | 6:116795040 | A | G | $8.26 \times 10^{-10}$ | 0.02 | intergenic | *CALHM6* | | |

| | | | | | | | | | |
|---|---|---|---|---|---|---|---|---|---|
| rs9400917 | 6:116794335 | A | G | $1.03 \times 10^{-9}$ | 0.02 | intergenic | *CALHM6* | | |
| rs34900657 | 6:116808418 | G | T | $1.07 \times 10^{-9}$ | 0.02 | intergenic | *CALHM6* | *POD* | |
| rs6925168 | 6:116799297 | A | G | $1.39 \times 10^{-9}$ | 0.01 | intergenic | *CALHM6* | | |
| rs4945556 | 6:116785195 | G | A | $1.64 \times 10^{-9}$ | 0.01 | downstream | *CALHM6* | | |
| rs7759971 | 6:135746884 | T | C | $3.11 \times 10^{-14}$ | 0.52 | intronic | AHI1 | | CD34, CLP, CMP, HSC, MPP |
| rs6928977 | 6:135626348 | T | G | $1.99 \times 10^{-13}$ | 0.09 | intronic | AHI1 | | |
| rs11154801 | 6:135739355 | A | C | $1.97 \times 10^{-13}$ | 0.08 | intronic | AHI1 | | CD34, CMP, HSC, LMPP, MEP, MPP |
| rs2614257 | 6:135676404 | C | T | $3.95 \times 10^{-13}$ | 0.04 | intronic | AHI1 | | CD34, CMP, MEP, MPP |
| rs4896143 | 6:135635100 | G | C | $5.95 \times 10^{-13}$ | 0.03 | intronic | AHI1 | | CD8, LMPP, MEP |
| rs2614266 | 6:135716532 | A | T | $7.74 \times 10^{-13}$ | 0.02 | intronic | AHI1 | | MPP |
| rs6935146 | 6:135627369 | T | C | $8.40 \times 10^{-13}$ | 0.02 | intronic | AHI1 | | *CMP* |
| rs6914831 | 6:135639644 | C | T | $9.90 \times 10^{-13}$ | 0.02 | intronic | AHI1 | | |
| rs6931735 | 6:135624811 | G | A | $1.13 \times 10^{-12}$ | 0.02 | intronic | AHI1 | | |
| rs6570001 | 6:135651721 | G | C | $1.28 \times 10^{-12}$ | 0.01 | intronic | AHI1 | | B |
| rs7759677 | 6:135909796 | C | T | $1.39 \times 10^{-12}$ | 0.01 | intergenic | AHI1 | PT1,PT2,PT3,PT | |
| rs761357 | 6:135902599 | T | A | $1.55 \times 10^{-12}$ | 0.01 | intergenic | AHI1 | | |
| rs6908428 | 6:135793706 | G | A | $2.10 \times 10^{-12}$ | 0.01 | intronic | AHI1 | | CMP |
| rs2179780 | 6:135650266 | A | G | $2.49 \times 10^{-12}$ | 0.01 | intronic | AHI1 | | MEP |
| rs11154806 | 6:135873721 | G | T | $2.64 \times 10^{-12}$ | 0.01 | intergenic | AHI1 | | |
| rs2246943 | 6:135691516 | A | T | $3.02 \times 10^{-12}$ | 0.01 | intronic | AHI1 | | |
| rs7772681 | 6:135649013 | T | C | $2.91 \times 10^{-12}$ | 0.01 | intronic | AHI1 | | |
| rs3827780 | 6:135709760 | G | A | $3.49 \times 10^{-12}$ | 0.01 | intronic | AHI1 | | |
| rs2746432 | 6:135696597 | C | T | $3.49 \times 10^{-12}$ | 0.01 | intronic | AHI1 | | |
| rs2327613 | 6:135662372 | T | C | $3.57 \times 10^{-12}$ | .005 | intronic | AHI1 | | |
| rs2614255 | 6:135663581 | T | C | $3.94 \times 10^{-12}$ | .004 | intronic | AHI1 | | |
| rs2746431 | 6:135687201 | C | T | $4.14 \times 10^{-12}$ | .004 | intronic | AHI1 | | |
| rs2246852 | 6:135691792 | G | A | $4.28 \times 10^{-12}$ | .004 | intronic | AHI1 | | |
| rs13197384 | 6:135818897 | A | C | $5.15 \times 10^{-12}$ | .004 | UTR5 | AHI1 | POD,PEC,MESFIB,ENDO,PT1, PT2,PT3,PT,LH,DCT,CNT,PC, ICA,ICB,LEUK | B, CD4, CD8, CD34, CLP, CMP, ERY, GMP, HSC, LMPP, MEP, MONO, MPP, NK |
| rs2614276 | 6:135681704 | T | C | $5.22 \times 10^{-12}$ | .004 | intronic | AHI1 | | CMP |
| rs12206850 | 6:135797808 | C | T | $5.76 \times 10^{-12}$ | .003 | intronic | AHI1 | | |
| rs9385726 | 6:135836962 | T | C | $6.49 \times 10^{-12}$ | .003 | intronic | LINC00271 | | |
| rs9647635 | 6:135841056 | C | A | $6.49 \times 10^{-12}$ | .003 | intergenic | LINC00271 | | |
| rs2256318* | 6:31381519 | A | G | $9.71 \times 10^{-18}$ | .270 | intronic | *MICA* | | |
| rs2256026* | 6:31379141 | G | C | $1.28 \times 10^{-17}$ | .206 | intronic | *MICA* | | |
| rs2256328* | 6:31381637 | G | C | $1.71 \times 10^{-17}$ | .155 | intronic | *MICA* | | |
| rs75918478* | 6:31453053 | A | G | $1.72 \times 10^{-17}$ | .154 | intergenic | *MICB* | | |
| rs2857282* | 6:31380807 | A | T | $2.87 \times 10^{-17}$ | .093 | intronic | *MICA* | | |
| rs2853982* | 6:31378751 | G | A | $5.64 \times 10^{-17}$ | .048 | intronic | *MICA* | | |
| rs2256028* | 6:31379198 | A | C | $6.33 \times 10^{-17}$ | .043 | intronic | *MICA* | | |
| rs6478108 | 9:117558703 | T | C | 7.48 x 10-11 | 0.25 | intronic | TNFSF15 | | |
| rs7848647 | 9:117569046 | C | T | 8.34 x 10-11 | 0.23 | upstream | TNFSF15 | PT1,PT2,PT3,PT,CNT,PC, ICB,LEUK | B, CD4, CMP, GMP, MPP |

| | | | | | | | | | |
|---|---|---|---|---|---|---|---|---|---|
| rs10817678 | 9:117579457 | A | G | 9.86 x 10-11 | 0.19 | intergenic | TNFSF15 | | |
| rs4263839 | 9:117566440 | G | A | 1.41 x 10-10 | 0.14 | intronic | TNFSF15 | | |
| rs6478109 | 9:117568766 | G | A | 1.87 x 10-10 | 0.10 | upstream | TNFSF15 | PEC,ENDO,PT1,PT2,PT3,PT,LH ,DCT,CNT,PC,ICA,ICB,LEUK | B, CD4, CD34, CMP, GMP, HSC, LMPP, MPP |
| rs4366152 | 9:117564875 | C | T | 2.21 x 10-10 | 0.09 | intronic | TNFSF15 | | |

Excluding HLA until after conditional analysis; PIP = posterior inclusion probability, EA = effect allele, NEA = non-effect allele

**Kidney cell type codes:** POD = podocyte, PEC = parietal epithelial cells, MES-FIB = mesangial and fibroblasts, ENDO = endothelial, PT(1-3) = proximal tubule, PT-KIM1P = proximal tubule with KIM1+ expression, LH = loop of Henle, DCT = distal convoluted tubule, CNT = connecting tubule, PC = principal cells, ICA = Type A intercalated cells, ICB = Type B intercalated cells, LEUK = leukocytes

**Immune cell type codes**: B = CD19+CD20+ B. CD4T, CD8T = CD4+ and CD8+ T, CD34 = CD34+ bone marrow and cord blood, CLP = common lymphoid progenitor, CMP = common myeloid progenitor, Ery = CD71+GPA+ erythroblast, GMP = granulocyte macrophage progenitor, HSC = hematopoietic stem, LMPP = lymphoid-primed multipotent progenitor, MEP = megakaryocyte erythroid progenitor, Mono = CD14+ monocyte, MPP = multipotent progenitor, NK = CD56+ natural killer T

**Table S8. Samples used for HLA fine-mapping analysis.**

| GWAS Cohort | Ancestry | n cases | n controls | n total |
|---|---|---|---|---|
| Nephrovir/EU | European | 313 | 2,508 | 2,821 |
| US Cohort | European | 361 | 4,309 | 4,670 |
| | European Total | 674 | 6817 | 7491 |
| US Cohort | African | 65 | 7,335 | 7,400 |
| Nephrovir/EU | African | 44 | 179 | 223 |
| | African Total | 109 | 7514 | 7623 |
| US Cohort | South Asian | 31 | 338 | 369 |
| Indian | South Asian | 162 | 98 | 260 |
| | South Asian Total | 193 | 436 | 629 |
| US Cohort | East Asian | 16 | 439 | 455 |
| Maghrebian | Maghrebian | 55 | 228 | 283 |
| Admixed American | Admixed American | 98 | 13,248 | 13,346 |
| Total | | 1,145 | 28,682 | 29,827 |

**Table S9. Genome-wide and suggestive loci from ancestry-specific HLA logistic regression and omnibus test.**

| Ancestry[a] | Logistic regression[b] | | | Omnibus test | |
|---|---|---|---|---|---|
| | SNP | P-value | OR [95% CI] | Amino Acid ID[c] | P-value |
| European | rs28755181 | $3.49 \times 10^{-51}$ | 3.35 [2.86 – 3.92] | AA_DQA1_47_32609219 | $6.81 \times 10^{-61}$ |
| African | rs1264705 | $3.03 \times 10^{-7}$ | 4.64 [2.58 – 8.34] | AA_DQB1_89_32632585 | $1.41 \times 10^{-8}$ |
| South Asian | HLA_B*44:03 | $9.42 \times 10^{-12}$ | 2.83 [2.12 - 3.77] | AA_B_199_31323321 | $5.64 \times 10^{-10}$ |
| Admixed-American | - | - | - | AA_DQA1_52_32609234 | $3.79 \times 10^{-11}$ |
| Maghrebian | rs9273344 | $2.543 \times 10^{-6}$ | 0.18 [0.09, 0.37] | AA_DQB1_-6_32634305 | $4.03 \times 10^{-5}$ |

OR [95% CI] = Odds ratio and 95% confidence interval.
[a] No significant associations in the East Asian analysis.
[b] Test included SNPs and classical HLA alleles.
[c] Amino acid (AA) _ gene _ AA position _ genomic position

**Supplemental note 1:**
The strongest association was rs2856696, between *HLA-DRB1* and *HLA-DQA1* ($P$ = 2.31 x $10^{-68}$). The strongest classical HLA allele association was at *DQA1*02* ($P$ = 2.79 X $10^{-59}$). Stepwise conditional analysis identified an independent association near *HLA-DQB1* (rs9273479; $P$ = 9.97 X $10^{-34}$).

**Table S10. Multi-population HLA.** Results from HLA logistic association test. Top overall association and top classical HLA allele both reported. Results after conditioning on rs2856696 also shown.

| | SNP | EA | P-value | OR | Closest Gene |
|---|---|---|---|---|---|
| Logistic | | | | | |
| | rs2856696 | T | 2.31 x $10^{-68}$ | 2.80 | *HLA-DRB1, HLA-DQA1* |
| | HLA_DQA1*02 | T | 2.79 x $10^{-59}$ | 2.49 | *HLA-DQA1* |
| Conditional Logistic | | | | | |
| | rs9273479 | T | 9.97 x $10^{-34}$ | 0.49 | *HLA-DQB1* |
| | HLA_DQA1*01 | T | 3.92 x $10^{-29}$ | 0.53 | *HLA-DQA1* |

**Figure S8.** Logistic regression results for HLA-DQA1 (A) and HLA-DQB1 (B) 4-digit alleles in multi-population analysis. Associations reaching genome-wide significance are indicated with a *

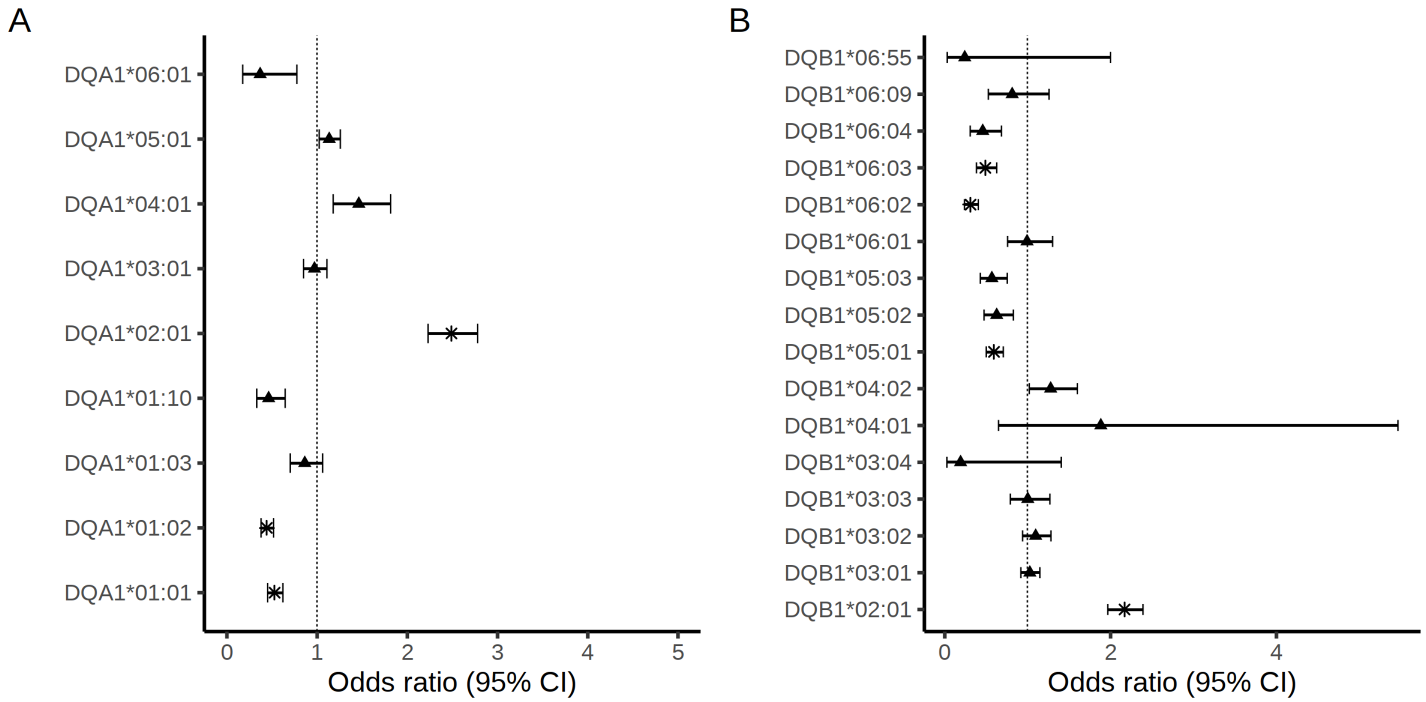


**Table S11. HLA amino acid omnibus test results.** Omnibus p-value is from an ANOVA comparing logistic models with and without amino acid position. Amino acid associations from logistic regression including all amino acid residues at the given position.

| HLA gene & Position | Omnibus P-value | Amino acid | OR [95% CI] | P-value |
|---|---|---|---|---|
| DQA1_47 | $7.73 \times 10^{-83}$ | | | |
| | | cysteine | 1.97 [1.75, 2.23] | $5.47 \times 10^{-28}$ |
| | | lysine | 3.62 [3.17, 4.14] | $5.70 \times 10^{-80}$ |
| | | glutamine | 1.72 [1.47, 2.00] | $7.63 \times 10^{-12}$ |
| | | arginine | ref | |
| DQA1_52 | $1.14 \times 10^{-82}$ | | | |
| | | serine | 0.53 [0.47, 0.59] | $1.00 \times 10^{-28}$ |
| | | histidine | 1.92 [1.70, 2.16] | $2.55 \times 10^{-27}$ |
| | | arginine | REF | |
| DQB1_26* | $3.22 \times 10^{-13}$ | | | |
| | | tyrosine | 0.90 [0.81, 1.00] | 0.06 |
| | | glycine | 0.64 [0.60, 0.73] | $4.75 \times 10^{-12}$ |
| | | leucine | ref | |

* Top association after conditioning on DQA1_47 and DQA1_52

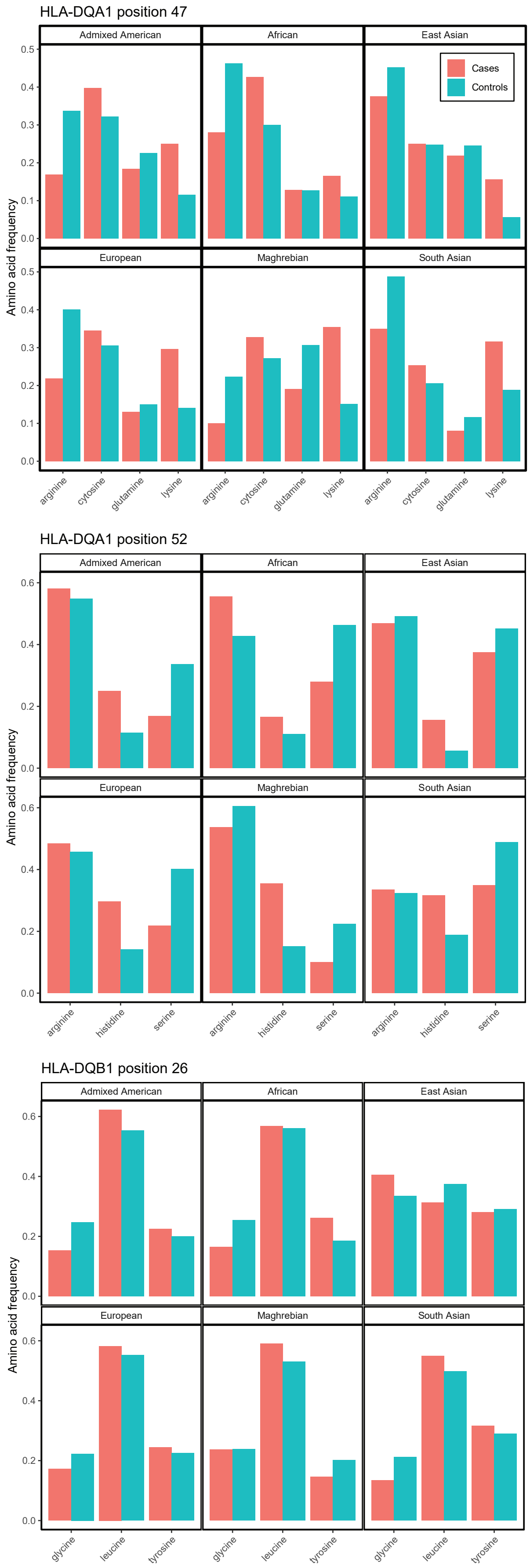


**Figure S9. Case/control frequency of amino acid residues at positions 47 and 52 of HLA-DQA1 and 26 of HLA-DQB1.**

**Table S12**. Results of Polygenic Risk Score (PRS) analysis in European and NEPTUNE cohorts.

| | Count (%) or Median (IQR) | PRS (IQR) | Univariate P-value [c] | Multiple linear regression P-value [d] |
|---|---|---|---|---|
| **European** [a] | n=233[e] | 1.79 [0.97-2.35] | - | - |
| Sex | | | 0.07 | - |
| Male | 146 (63%) | 1.89 [0.98-2.43] | | |
| Female | 87 (37%) | 1.51 [0.90-2.14] | | |
| Steroid sensitivity | | | 0.17 | - |
| SDNS | 106 (45%) | 1.84 [1.13-2.44] | | |
| SSNS | 127 (55%) | 1.70 [0.91-2.19] | | |
| Age onset | 4.40 (3.09-7.20) | | $4.13 \times 10^{-4}$ | $1.49 \times 10^{-4}$ |
| **NEPTUNE** | N=165 | 0.82 [0.50-1.09] | - | - |
| Sex | | | 0.35 | - |
| Male | 97 (59%) | 0.82 [0.43-1.08] | | |
| Female | 68 (41%) | 0.82 [0.61-1.10] | | |
| Histology | | | 0.37 | - |
| FSGS | 50 (30%) | 0.80 [0.48-1.03] | | |
| MCD | 81 (49%) | 0.83 [0.50-1.12] | | |
| Non-biopsy | 34 (21%) | 0.81 [0.61-0.95] | | |
| Age of onset | 5.00 (3.00-11.00) | | 0.02 | 0.003 |
| Ancestry [b] | | | 0.02 | - |
| African | 58 (35%) | 0.86 [0.64-1.09] | | |
| Admixed American | 32 (19%) | 0.75 [0.47-0.90] | | |
| East Asian | 6 (4%) | 0.44 [0.08-0.60] | | |
| EUR | 59 (36%) | 0.86 [0.66-1.15] | | |
| SAS | 10 (6%) | 0.55 [0.44-0.88] | | |

[a] European cohort from Paris
[b] Predicted ancestry based on genotype data and 1000 genomes reference panel
[c] Wilcoxon test for binary traits, Kruskal-Wallis for categorical traits, univariate linear regression for continuous traits
[d] European (Paris cohort) adjusted for sex, steroid sensitivity and 4 genetic principal components. NEPTUNE adjusted for sex, histology and 4 genetic principal components
[e] 233 out of a total 311samples had available demographic information

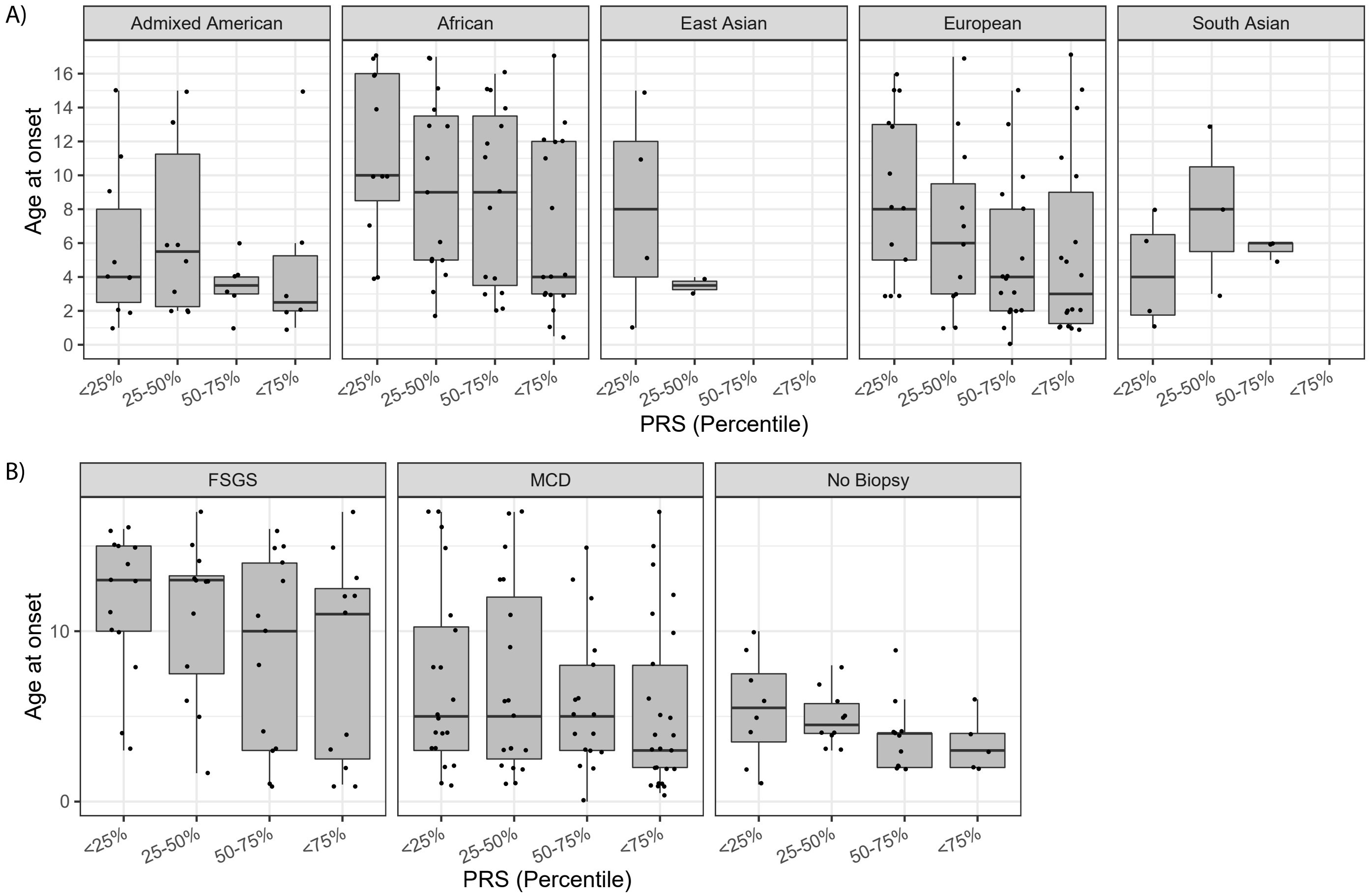


**Figure S10. Association between PRS quartiles and age of onset in the NEPTUNE cohort. A)** Stratified by genotype-predicted ancestry. **B)** By histology.

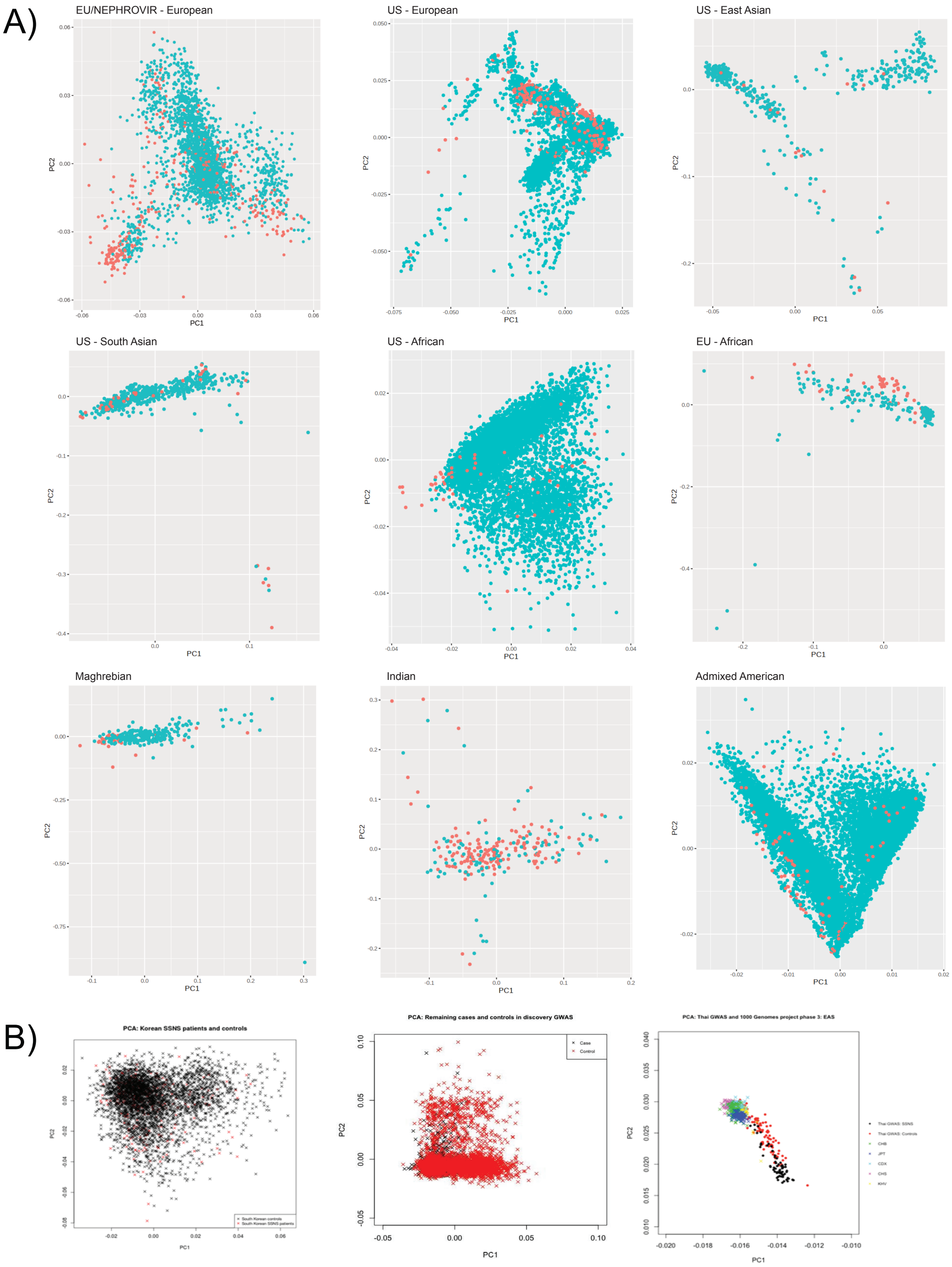


**Figure S11. PCA (PC1 vs. PC2) of dataset GWAS cases and controls**.
**A)** Data for which genotype-level data were available. Cases are indicated in red; controls are indicated in blue.
**B)** Data with only summary statistics were available.

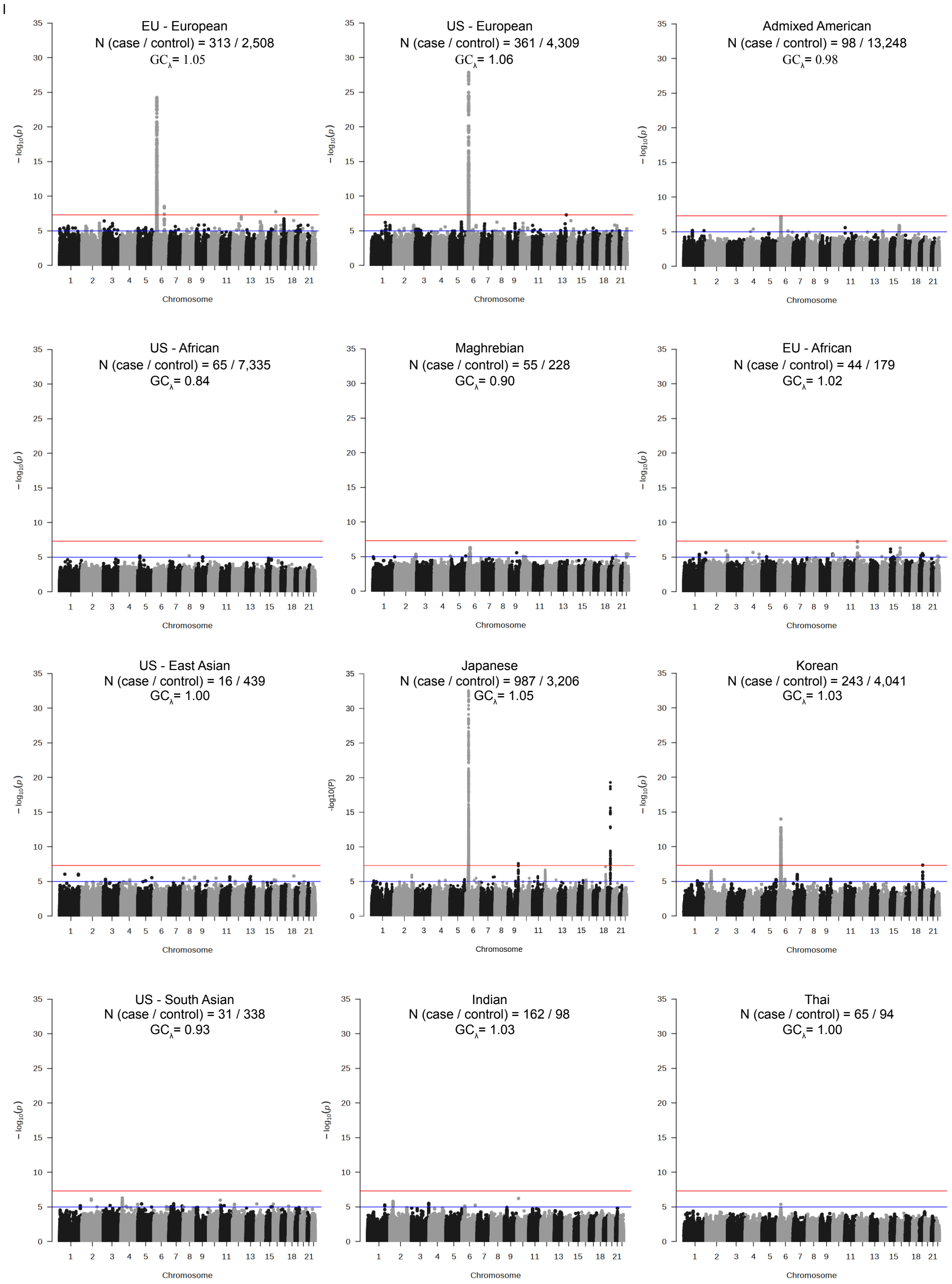


**Figure S12. Manhattan plot of GWAS by cohort.**

Red line indicated genome-wide significance threshold ($5 \times 10^{-8}$); blue line is suggestive significance ($1 \times 10^{-5}$). Number of cases and controls and genomic controls lambda are reported for each dataset.

**Table S13: Genome-wide significant loci from individual GWAS**

All SNP are > 1Mb from each other with $r^2 < 0.1$. We found no genome-wide significant associations in the Thai, EU – African, Maghrebian, Indian, Admixed American, US – East Asian and US – South Asian datasets.

| Dataset | Top SNP | Position (hg19) | Nearest Gene | EA | NEA | OR [95% CI] | P |
|---|---|---|---|---|---|---|---|
| EU - European | rs9275205 | 6:32657560 | *HLA-DQB1* | C | T | 3.13 [2.52, 3.88] | $5.69 \times 10^{-25}$ |
| EU - European | rs2637681 | 6:116769845 | *DSE* | G | T | 0.54 [0.44, 0.66] | $3.01 \times 10^{-9}$ |
| EU - European | rs17185137 | 16:26809795 | *C16orf82* | G | A | 3.03 [2.06, 4.46] | $1.78 \times 10^{-8}$ |
| US - European | rs1694129 | 6:32637743 | *HLA-DQB1* | T | G | 3.20 [2.61, 3.93] | $1.38 \times 10^{-28}$ |
| Japanese | rs6901541 | 32442261 | *HLA-DRA* | C | T | 2.49 [2.15, 2.89] | $2.81 \times 10^{-33}$ |
| Japanese | rs56117924 | 36334182 | *NPHS1* | A | G | 1.90 [1.66, 2.18] | $4.95 \times 10^{-20}$ |
| Japanese | rs117576077 | 31906997 | *C2* | T | C | 0.13 [0.07, 0.23] | $1.85 \times 10^{-11}$ |
| Japanese | rs3117032 | 33088592 | *HLA-DPB2* | T | C | 0.40 [0.30, 0.53] | $1.88 \times 10^{-10}$ |
| Japanese | rs3095273 | 29566369 | *GABBR1* | A | G | 0.44 [0.34, 0.58] | $6.45 \times 10^{-9}$ |
| Japanese | rs6478109 | 117568766 | *TNFSF15* | A | G | 0.72 [0.64, 0.81] | $2.55 \times 10^{-8}$ |
| Korean | rs9272518 | 32606446 | *HLA-DQA1* | C | G | 2.63 [2.06, 3.35] | $1.05 \times 10^{-14}$ |
| Korean | rs412175 | 36342103 | *NPHS1* | C | T | 1.83 [1.47, 2.27] | $4.66 \times 10^{-8}$ |

**The Research Consortium on Genetics of Childhood Idiopathic Nephrotic Syndrome in Japan**

Yoshinori Araki[1], Yoshinobu Nagaoka[1], Takayuki Okamoto[2], Yasuyuki Sato[2], Asako Hayashi[2], Toshiyuki Takahashi[2], Hayato Aoyagi[3], Michihiko Ueno[4], Masanori Nakanishi[5], Nariaki Toita[6], Kimiaki Uetake[7], Norio Kobayashi[8], Shoji Fujita[9], Kazushi Tsuruga[10], Naonori Kumagai[11, 12], Hiroki Kudo[11], Eriko Tanaka[13, 14], Tae Omori[15], Mari Okada[16], Yoshiho Hatai[17], Tomohiro Udagawa[18, 19], Yaeko Motoyoshi[20], Koichi Kamei[21], Masao Ogura[21], Mai Sato[21], Yuji Kano[21, 22], Motoshi Hattori[23], Kenichiro Miura[23], Yutaka Harita[24], Shoichiro Kanda[24], Emi Sawanobori[25], Anna Kobayashi[25], Manabu Kojika[26], Yoko Ohwada[27, 28], Kunimasa Yan[29], Hiroshi Hataya[30], Riku Hamada, Chikako Terano[30], Ryoko Harada[30], Yuko Hamasaki[31], Junya Hashimoto[31], Kenji Ishikura[32], Shuichi Ito[33], Hiroyuki Machida[33], Aya Inaba[33], Takeshi Matsuyama[34], Miwa Goto[35], Masaki Shimizu[36], Kazuhide Ohta[37], Yohei Ikezumi[38, 39], Takeshi Yamada[38], Toshiaki Suzuki[40], Soichi Tamamura[41], Yukiko Mori[41], Yoshihiko Hidaka[42], Daisuke Matsuoka[42], Tatsuya Kinoshita[43], Shunsuke Noda[44], Masashi Kitahara[45], Naoya Fujita[46], Satoshi Hibino[46], Kandai Nozu[47], Tomoko Horinouchi[47], Tomohiko Yamamura[47], China Nagano[47], Shogo Minamikawa[47, 48], Keita Nakanishi[47, 49], Junya Fujimura[47, 50], Nana Sakakibara[47], Yuya Aoto[47], Shinya Ishiko[47], Kazumoto Iijima[51, 52], Ryojiro Tanaka[51], Hiroshi Kaito[51], Kyoko Kanda[51, 53], Yosuke Inaguma[51], Yuya Hashimura[54], Shingo Ishimori[55, 56], Naohiro Kamiyoshi[57], Takayuki Shibano[58], Yasuhiro Takeshima[58], Rika Fujimaru[59], Hiroaki Ueda[59], Akira Ashida[60], Hideki Matsumura[60], Takuo Kubota[61], Taichi Kitaoka[61, 62], Yusuke Okuda[63, 64], Toshihiro Sawai[63], Tomoyuki Sakai[63], Yuko Shima[65], Taketsugu Hama[65], Mikiya Fujieda[66], Masayuki Ishihara[66], Shigeru Itoh[67], Takuma Iwaki[68], Maki Shimizu[69], Koji Nagatani[70], Shoji Kagami[71], Maki Urushihara[71], Yoshitsugu Kaku[72], Manao Nishimura[72], Miwa Yoshino[72], Ken Hatae[73], Maiko Hinokiyama[73], Rie Kuroki[73], Yasufumi Ohtsuka[74], Masafumi Oka[74], Shinji Nishimura[75], Tadashi Sato[76], Seiji Tanaka[77], Ayuko Zaitsu[77], Hitoshi Nakazato[78], Hiroshi Tamura[78], Koichi Nakanishi[79]

1. Department of Pediatrics, National Hospital Organization Hokkaido Medical Center, Sapporo, Japan
2. Department of Pediatrics, Hokkaido University Hospital, Sapporo, Japan
3. Department of Pediatrics, Obihiro Kyokai Hospital, Obihiro, Japan
4. Department of Pediatrics, Nikko Memorial Hospital, Muroran, Japan
5. Department of Pediatrics, Kushiro Red Cross Hospital, Kushiro, Japan
6. Department of Pediatrics, Sapporo Kosei Hospital, Sapporo, Japan
7. Department of Pediatrics, Obihiro Kosei Hospital, Obihiro, Japan
8. Department of Pediatrics, Oji General Hospital, Tomakomai, Japan
9. Department of Pediatrics, Hakodate Goryoukaku Hospital, Hakodate, Japan
10. Department of Pediatrics, Hirosaki University Hospital, Hirosaki, Japan
11. Department of Pediatrics, Tohoku University Graduate School of Medicine, Sendai, Japan
12. Present address: Department of Pediatrics, Fujita Health University, Toyoake, Japan
13. Department of Pediatrics and Developmental biology, Tokyo Medical and Dental University, Tokyo, Japan
14. Present address: Department of Pediatrics, Kyorin University Hospital, Tokyo, Japan
15. Department of Peditrics, Tokyo Metropolitan Bokutoh Hospital, Tokyo, Japan
16. Musashino Red Cross Hospital, Musashino, Japan
17. Tokyo Bay Urayasu-Ichikawa Medical Center, Urayasu, Japan
18. Tsuchiura Kyodo General Hospital, Tsuchiura, Japan
19. Present address: Department of Pediatrics and Developmental biology, Tokyo Medical and Dental University, Tokyo, Japan
20. Department of Pediatrics, Tokyo Kita Medical Center, Tokyo, Japan
21. Division of Nephrology and Rheumatology, National Center for Child Health and Development, Tokyo, Japan
22. Present address: Department of Pediatrics, Dokkyo Medical University School of Medicine, Mibu, Japan
23. Department of Pediatric Nephrology, Tokyo Women's Medical University, Tokyo, Japan

24. Department of Pediatrics, The University of Tokyo Hospital, Tokyo, Japan
25. Department of Pediatrics, Faculty of Medicine, University of Yamanashi, Chuo, Japan
26. Department of Pediatrics, Fujiyoshida Manucipal Hospital, Fujiyoshida, Japan
27. Department of Pediatrics, Dokkyo Medical University School of Medicine, Mibu, Japan
28. Department of Pediatrics,Tochigi Medical Center Shimostuga, Tochigi, Japan
29. Department of Pediatrics, Kyorin University Hospital, Tokyo, Japan
30. Department of Nephrology, Tokyo Metropolitan Children’s Medical Center, Tokyo, Japan
31. Department of Nephrology, Toho University Faculty of Medicine, Tokyo, Japan
32. Department of Pediatrics, Kitasato Univeristy School of Medicine, Sagamihara, Japan
33. Department of Peditrics, Yokohama City University, Yokohama, Japan
34. Department of Pediatrics, Fussa Hospital, Tokyo, Japan
35. Department of Pediatrics, National Hospital Organization Kofu National Hospital, Kofu, Japan
36. Department of Pediatrics, Kanazawa University Hospital, Kanazawa, Japan
37. Department of Pediatrics, Kanazawa Medical Center, Kanazawa, Japan
38. Department of Pediatrics, Niigata University Medical & Dental Hospital, Niigata, Japan
39. Present address: Department of Pediatrics, Fujita Health University, Toyoake, Japan
40. Department of Pediatrics, National Hospital Organization Niigata National Hospital, Niigata
41. Department of Pediatrics, Japanese Red Cross Fukui Hospital, Fukui, Japan
42. Department of Pediatrics, Shinshu University Hospital, Matsumoto, Japan
43. Department of Pediatrics, Ina Central Hospital, Ina, Japan
44. Department of Pediatrics, Nagano Red Cross Hospital, Nagano, Japan
45. Department of Pediatrics, Matsumoto Medical Center, Matsumoto, Japan
46. Department of Pediatric Nephrology, Aichi Children's Health And Medical Center, Obu, Japan
47. Department of Pediatrics, Kobe University Graduate School of Medicine, Kobe, Japan
48. Present address: Department of General Medicine, Hyogo Prefectural Kobe Children's Hospital, Kobe, Japan
49. Present address: Department of Pediatrics, Saiseikai Hyogoken Hospital, Kobe, Japan
50. Present address: Department of Pediatrics, Kakogawa Central City Hospital, Kakogawa, Japan
51. Department of Nephrology, Hyogo Prefectural Kobe Children's Hospital, Kobe, Japan
52. Department of Advanced Pediatric Medicine, Kobe University Graduate School of Medicine, Kobe, Japan
53. Department of Pediatrics, National Hospital Organization Kobe Medical Center
54. Department of Pediatrics, Takatsuki General Hospital, Takatsuki, Japan
55. Department of Pediatrics, Kakogawa Central City Hospital, Kakogawa, Japan
56. Present address: Department of Pediatrics, Takatsuki General Hospital, Takatsuki, Japan
57. Department of Pediatrics, Himeji Red Cross Hospital, Himeji, Japan
58. Department of Pediatrics, Hyogo College of Medicine, Nishinomiya, Japan
59. Department of Pediatrics, Osaka City General Hospital, Osaka, Japan
60. Department of Pediatrics, Osaka Medical College, Takatsuki, Japan
61. Department of Pediatrics, Osaka Univeristy Graduate School of Medicine, Suita, Japan
62. Present address: Department of Pediatrics, Yodogawa Children Hospital, Osaka, Japan
63. Department of Pediatrics, Shiga University of Medical Science, Otsu, Japan
64. Present address: Department of Pediatrics, Kitasato Univeristy School of Medicine, Sagamihara, Japan
65. Department of Pediatrics, Wakayama Medical University, Wakayama, Japan
66. Department of Pediatrics, Kochi Medical School, Kochi University, Nankoku, Japan
67. Department of Pediatrics, Kagawa Prefecture Central Hospital, Takamatsu, Japan
68. Department of Pediatrics, Faculty of Medicine, Kagawa University, Kagawa, Japan
69. Department of Pediatrics, Faculty of Medicine, Kagawa University, Kagawa, Japan
70. Department of Pediatrics, Uwajima City Hospital, Uwajima, Japan
71. Department of Pediatrics, Institute of Biomedical Sciences, Tokushima University Graduate School, Tokushima, Japan
72. Department of Nephrology, Fukuoka Children’s Hospital, Fukuoka, Japan
73. Department of Pediatrics, Japanese Red Cross Fukuoka Hospital, Fukuoka, Japan
74. Department of Pediatrics, Faculty of Medicine, Saga University, Saga, Japan

75. Department of Pediatrics, Saga-ken Medical Centre Koseikan, Saga, Japan
76. Department of Pediatrics, National Hospital Organization Ureshino Medical Center, Ureshino, Japan
77. Department of Pediatrics and Child Health, Kurume University School of Medicine, Kurume, Japan
78. Department of Pediatrics, Faculty of Life Sciences, Kumamoto University, Kumamoto, Japan
79. Department of Child Health and Welfare (Pediatrics), Graduate School of Medicine, University of the Ryukyus, Nishihara, Japan

**Korean Consortium of Hereditary Renal Diseases in Children**

Min Hyun Cho[1], Tae-Sun Ha[2], Hee Gyung Kang[3], Il-Soo Ha[3], Ji Hyun Kim[3], Peong Gang Park[3], Kyoung Hee Han[4], Eun Mi Yang[5], Myung Hyun Cho[6], Hae Il Cheong[6]

1. Department of Pediatrics, Kyungpook National University, School of Medicine, Daegu, Korea
2. Department of Pediatrics, Chungbuk National University College of Medicine, Cheongju, Korea
3. Department of Pediatrics, Seoul National University Children's Hospital, Seoul, Korea
4. Department of Pediatrics, Jeju National University School of Medicine, Jeju, Korea
5. Department of Pediatrics, Chonnam National University Children's Hospital, Gwangju, Korea
6. Department of Pediatrics, Hallym University Sacred Heart Hospital, Anyang, Korea

**Thailand Team**

Prayong Vachvanichsanong[1], Kwanchai Pirojsakul[2]

1. Department of Pediatrics, Faculty of Medicine, Prince of Songkla University, Hat-Yai, Songkhla, Thailand
2. Department of Pediatrics, Faculty of Medicine, Ramathibodi Hospital Mahidol University, Bangkok, Thailand

**ACKNOWLEDGMENTS:**

The authors wish to thank Seong Kyu Han, PhD (Boston Children's Hospital and Harvard Medical School) for his assistance in creating Figure 1.

- The Nephrotic Syndrome Study Network (NEPTUNE) is part of the Rare Diseases Clinical Research Network (RDCRN), which is funded by the National Institutes of Health (NIH) and led by the National Center for Advancing Translational Sciences (NCATS) through its Division of Rare Diseases Research Innovation (DRDRI). NEPTUNE is funded under grant number U54DK083912 as a collaboration between NCATS and the National Institute of Diabetes and Digestive and Kidney Diseases (NIDDK). Additional funding and/or programmatic support is provided by the University of Michigan, NephCure Kidney International and the Halpin Foundation. RDCRN consortia are supported by the RDCRN Data Management and Coordinating Center (DMCC), funded by NCATS and the National Institute of Neurological Disorders and Stroke (NINDS) under U2CTR002818.

- This study was funded by European Research Council grant ERC-2012- ADG_20120314 (grant agreement 322947) and Agence Nationale pour la Recherche "Genetransnephrose" grant ANR-16-CE17-004-01.

- MGS is supported by NIH grants R01DK119380, RC2DK122397, 2U54DK083912 and a gift from The Pura Vida Kidney Foundation
